# Between- and Within-Person Coupling of Nocturnal Heart Rate and Glucose: Retrospective, Real-World Observational Study of 105,196 Nights From 2648 Adults Using Ultrahuman Ring and Continuous Glucose Monitoring

**DOI:** 10.64898/2026.03.04.26347496

**Authors:** Nihav Dhawale, Dhruv Gandhi, Hans-Peter Kubis, Matthew Driller, Michael P. Snyder, Tao Wang, Aditi Bhattacharya

## Abstract

**Background:** Higher resting or sleeping heart rate (HR) has been associated with poorer glucose metabolism in adults without diabetes. However, these associations were estimated across individuals, and within-person characterization of this co-regulation during naturalistic sleep in large free-living cohorts remains limited.

**Objective:** This study aimed to determine whether average sleeping HR, an indirect index of cardiac autonomic tone, is associated with nocturnal glucose within a participant and between participants, and whether night-to-night irregularity in sleep duration, sleep onset, and wake time is associated with nocturnal glycemia.

**Methods:** In this retrospective observational study, adults who concurrently wore an Ultrahuman Ring and a continuous glucose monitor between January 2024 and June 2025 were analyzed. After participant- and night-level filtering, the cohort comprised 2648 participants from 87 countries, contributing 105,196 paired nights. Each predictor was decomposed into within- and between-participant components by person-mean centering and entered a single linear mixed-effects model with a random intercept for participant, adjusted for age, body mass index (BMI), and sex. Outcomes were nocturnal mean glucose, nocturnal glucose coefficient of variation (CV), and nocturnal time in range (70-110 mg/dL). Per-participant slopes were fitted in participants with sustained wear (at least 30 nights spanning at least 60 days). Estimates were re-derived under trimming and robust-regression specifications and refitted within BMI and regional strata.

**Results:** Higher average sleeping HR was associated with higher nocturnal mean glucose both within participants (+5.7 mg/dL per 10 bpm, 95% CI 5.4-5.9) and between participants (+6.5 mg/dL, 95% CI 5.6-7.3; both *P*<.001). Within participants, higher sleeping HR was also associated with higher glucose CV (+1.37 percentage points, 95% CI 1.30-1.43) and lower time in range (−6.3 percentage points, 95% CI −6.7 to −6.0). Of the 911 sustained-wear participants, 701 (76.9%, 95% CI 74.1%-79.6%) had a positive individual slope (median of +5.2 mg/dL per 10 bpm). The association survived in every BMI stratum and in all 5 regional strata. Sleep-duration irregularity was associated with mean glucose between participants (+4.1 mg/dL per 30 minutes, 95% CI 2.6-5.7) but only weakly within them (+0.23 mg/dL). Within participants, a night 1 standard deviation above a participant’s own mean sleeping HR corresponded to 2.5 mg/dL higher nocturnal mean glucose.

**Conclusions:** Average sleeping HR was associated with glucose within individuals as well as between them, indicating that the relationship is not attributable solely to stable between-person differences and reflects, at least in part, night-to-night covariation. The association was modest, but discernible in naturalistic sleep across geographies. Sleep-duration irregularity was associated with nocturnal glycemia mainly between individuals.

## Introduction

Sleep plays a central role in maintaining physiological homeostasis, coordinating both autonomic and metabolic processes. The autonomic nervous system shifts systematically across the stages of sleep, which is reflected in cyclical alterations in heart rate (HR), heart rate variability (HRV), and other cardiovascular parameters. This has also been linked to the glycemic state of the body during sleep [1–6]. Experimental and epidemiological evidence shows that dyssynchrony in this nexus is reflected in reduced HRV or elevated resting HR. These markers have been associated with impaired insulin sensitivity [4,7], greater nocturnal glucose variability [2], and adverse metabolic outcomes over time [7–9]. Sleep is therefore a period in which autonomic function and glucose metabolism interact closely, with implications for long-term cardiometabolic health and noncommunicable diseases.

Night-to-night fluctuations in sleep duration, timing, and architecture may destabilize autonomic and metabolic regulation. Population-level studies show that sleep quality has declined in recent decades, with at least 30% of the American population reporting less than 6 hours of nightly sleep [10,11] and similar patterns reported globally [12,13]. Sleep also has a well-established connection with long-term glycemic control [14–16], with much of that literature establishing links in individuals at heightened risk of diabetes [12,15,17], cardiovascular disease and noncommunicable diseases [17–19], or sleep apnea [20], based on average sleeping HR or on glucose patterns separately rather than jointly.

Evidence from individuals not selected for disease is less abundant. The published record suggests that it is heavily dependent on study design. Studies of HR and glucose during sleep in nominally healthy adults have generally been small and conducted under laboratory or protocol conditions [2,21,22], which optimizes measurement precision at the cost of the behavioral variety of ordinary sleep. Free-living studies avoid that constraint but have typically involved small, young, or single-site cohorts; in one, 69 university students ate a standardized diet and underwent a night of sleep restriction [23]. Whether an association observed under controlled conditions persists under unconstrained, free-living ones therefore remains uncertain.

Nocturnal HR may reflect underlying metabolic demand and autonomic regulation of glycemia through vagal-sympathetic balance, which modulates HR, hepatic glucose output, and pancreatic hormone secretion [24–26]. Recent human studies have shown that sleep microarchitecture and autonomic state transitions are temporally coupled with changes in interstitial glucose [21,22]. Nocturnal glucose merits a refined examination in this context as a reflection of endogenous regulation, without activity, food, and arousal states that impact daytime glucose levels.

Cross-sectional comparisons cannot distinguish whether an association reflects stable differences between individuals or a state that varies within individuals from night to night; repeated-measures designs that separate within-person and between-person components can. Smart rings and continuous glucose monitors (CGMs) make this feasible. In this study we analyzed free-living data from 2648 participants contributing 105,196 nights of concurrent Ultrahuman Ring and CGM wear. We asked whether average sleeping HR, an indirect marker of autonomic tone, is associated with nocturnal glucose within a participant as well as between participants, and whether night-to-night irregularity in sleep duration, sleep onset, and wake time is related to nocturnal glycemia. Given the shared autonomic regulation of HR and glucose, we expected higher sleeping HR to accompany higher nocturnal glucose both between and within participants. Because irregular sleep may destabilize autonomic and metabolic regulation, we also expected greater night-to-night irregularity to accompany higher nocturnal glucose.

## Methods

### Study Design and Participants

This retrospective observational study used data from users of the Ultrahuman platform. Between January 2024 and June 2025, 5859 concurrent users of the Ultrahuman Ring and the Ultrahuman CGM platform (using the Abbott FreeStyle Libre 2, Pro, and 3 models [27]) were identified, contributing 227,860 nights of overlapping CGM and HR data.

Age, sex, and body mass index (BMI) were self-reported at onboarding; BMI could be updated by the user thereafter. Participants self-reported no history of metabolic or cardiovascular disease and no use of relevant medications.

### Ethical Considerations

#### Ethics Approval

This retrospective analysis of de-identified records from Ultrahuman device users involved no intervention and no contact with participants, so the authors determined that institutional ethics board approval was not required. The device-validation sub-studies (Multimedia Appendix 1) rest on their own bases. The Compumedics Somfit comparison was approved by the La Trobe University Human Ethics Committee (HEC22369) as part of a larger independent study. The continuous HR comparison used data provided by a third party with written consent [28].

#### Informed Consent

Users consented to the research use of their de-identified, aggregated data through the Ultrahuman terms of use [29] and privacy policy [30], accepted at onboarding and by continued use of the platform. Volunteers in the validation sub-studies consented to wearing 2 devices concurrently.

#### Privacy and Confidentiality

Direct identifiers were removed before analysis. Data handling complied with the General Data Protection Regulation and the Indian Digital Personal Data Protection Act of 2023.

#### Compensation Details

No compensation was provided to participants.

### Devices and Measurements

#### Ultrahuman Ring

The Ultrahuman Ring carries a photoplethysmography sensor, an accelerometer, and a skin temperature sensor, and outputs HR, motion, and skin temperature data every 5 minutes. These signals are used to derive the sleep metrics used here (sleep onset, sleep offset [wake time], and sleep duration) and the HR metrics (average sleeping HR, lowest sleeping HR, and sleep HRV). The ring’s HRV metric is the root mean square of successive differences.

Sleep duration refers to scored sleep, defined as the sleep period excluding wake after sleep onset.

Average sleeping HR is the mean HR across the sleep period and is the primary exposure, used as an indirect index of nocturnal cardiac autonomic state; where unqualified, sleeping HR refers to this measure. HR reflects net sympathovagal input to the sinoatrial node and is also influenced by nonautonomic factors, including body temperature and metabolic rate. Lowest sleeping HR is the minimum of that 5-minute series over the scored sleep period and was treated as a secondary measure, because it is a single nightly minimum.

#### Validation of Ring Sleep and Heart-Rate Measures

Ring-derived sleep timing and HR were compared against independent reference devices. Ring sleep metrics were compared with the Compumedics Somfit in a separately reported dataset [31]. Ring average sleeping HR was compared with the Somfit (Figure S1 in Multimedia Appendix 1), and the continuous overnight HR series with the US Food and Drug Administration (FDA)-cleared SleepImage ring [28] (Table S1 in Multimedia Appendix 1). Ring HRV was not compared against a reference and was treated as a secondary measure. Cohorts, night-selection criteria, and agreement statistics are reported under Device Validation in Multimedia Appendix 1. These comparisons characterize measurement properties only; no estimate in this paper is derived from them.

#### Continuous Glucose Monitoring and Nocturnal Glucose Metrics

CGM sampling intervals were 1, 5, or 15 minutes, depending on sensor model and operating mode. Glucose data were resampled to a nominal 15-minute interval by time-weighted linear interpolation, and gaps longer than 32 minutes were treated as missing. Data density was defined as the ratio of observed to expected data points within a period, where expected points were the period duration divided by the nominal sampling interval; sessions with data density below 80% were excluded.

Nocturnal glucose metrics were computed using Glucose360, an open-source Python framework made available prior to publication through collaboration with the Snyder Laboratory, Stanford University School of Medicine [32]. The 3 nocturnal glucose metrics were mean glucose, glucose coefficient of variation (CV), and time in range. The standard 14-day computation window [33,34] was replaced by the sleep window, defined by ring-derived sleep onset and offset, so that each glucose metric described the same night as the paired sleep and HR metrics.

Time in range was computed over 70 to 110 mg/dL, consistent with the platform’s Metabolic Score described previously [35]. This range is narrower than the consensus target range used in diabetes care, 70 to 180 mg/dL [34].

A 24-hour daily mean glucose was also computed for each calendar day. It was not an outcome in any analysis. It was used only as a plausibility bound in the inclusion criteria, to compare the cohort with published CGM reference cohorts, whose values are reported over 24 hours, and to compare countries with national diabetes prevalence.

#### Platform Composite Scores

Two proprietary composite scores were used as secondary variables and neither was the basis of any primary estimate.

The Ultrahuman Sleep Score (0-100; higher indicates better sleep) is a proprietary composite. Its contributors include sleep duration, sleep efficiency, number of awakenings, sleep staging, consistency of sleep onset and offset timing, sleeping HR, skin temperature, and movement during sleep. The consistency contributor scores how closely a night’s sleep onset and offset match the participant’s preceding nights and is undefined for a night with too little preceding history; this study used it only to exclude such nights.

The Ultrahuman Metabolic Score (0-100; higher indicates better glucose regulation) is computed from CGM data, using glucose CV, mean glucose, and time in range (70-110 mg/dL), as described previously [35]. It was computed over the sleep window (Night Metabolic Score) for all subgrouping, because the construct of interest was the overnight rather than the 24-hour state; the 24-hour version (Daily Metabolic Score) is reported in Table 1 only.

**Table 1.** Cohort characteristics (n=2c48 participants, 105,1Sc nights).

| Characteristic | Value |
| --- | --- |
| **Cohort** |  |
| Participants, n | 2648 |
| Nights, n | 105,196 |
| Nights per participant | 22 (13-44) |
| **Demographics** |  |
| Age, y | 42 (35-49) |
| BMI, kg/m<sup>2</sup> | 26.3 (23.7-29.8) |
| Male, n (%) | 1975 (74.6) |
| Female, n (%) | 673 (25.4) |
| **Geography (modal country)** |  |
| Countries, n | 87 |
| United States, n (%) | 781 (29.5) |
| India, n (%) | 707 (26.7) |
| United Kingdom, n (%) | 176 (6.6) |
| United Arab Emirates, n (%) | 171 (6.5) |
| Czechia, n (%) | 122 (4.6) |
| Germany, n (%) | 67 (2.5) |
| Other countries (81), n (%) | 624 (23.6) |
| **Sleep, per night** |  |
| Average sleeping HR, bpm | 67 (60-74) |
| Lowest sleeping HR, bpm | 57 (52-64) |
| Sleep HRV, ms | 36 (27-46) |
| Sleep duration, h | 7.0 (6.1-7.9) |
| Sleep Score | 79 (71-85) |
| **Nocturnal glucose, per night** |  |
| Mean glucose, mg/dL | 96 (85-111) |
| Glucose CV, % | 8.9 (6.1-12.9) |
| Time in range (70-110 mg/dL), % | 82 (42-100) |
| Night Metabolic Score | 86 (73-94) |
| Daily Metabolic Score | 76 (64-85) |

Beyond their use as secondary variables, the scores had 2 further roles. First, outlier fences on the Sleep Score and Night Metabolic Score were used to remove nights with implausible values, such as those arising from sensor displacement or nighttime travel. Each score summarized several signals and so flagged such nights efficiently. Because the Night Metabolic Score was computed from the same nocturnal glucose metrics that served as outcomes, filtering on it restricted the outcome range. Second, the Night Metabolic Score defined the extreme-group subcohorts, which were therefore reported descriptively.

### Analysis Dataset and Inclusion Criteria

Filters were applied to reduce measurement artifact, as shown in Figure 1. Analyses were restricted to participants aged 20 to 60 years with a BMI of 18 to 40 kg/m² who self-identified as male or female. Nights were retained if total sleep duration was 4 to 14 hours, to exclude fragmented or implausibly long sleep sessions.

**Figure 1.**
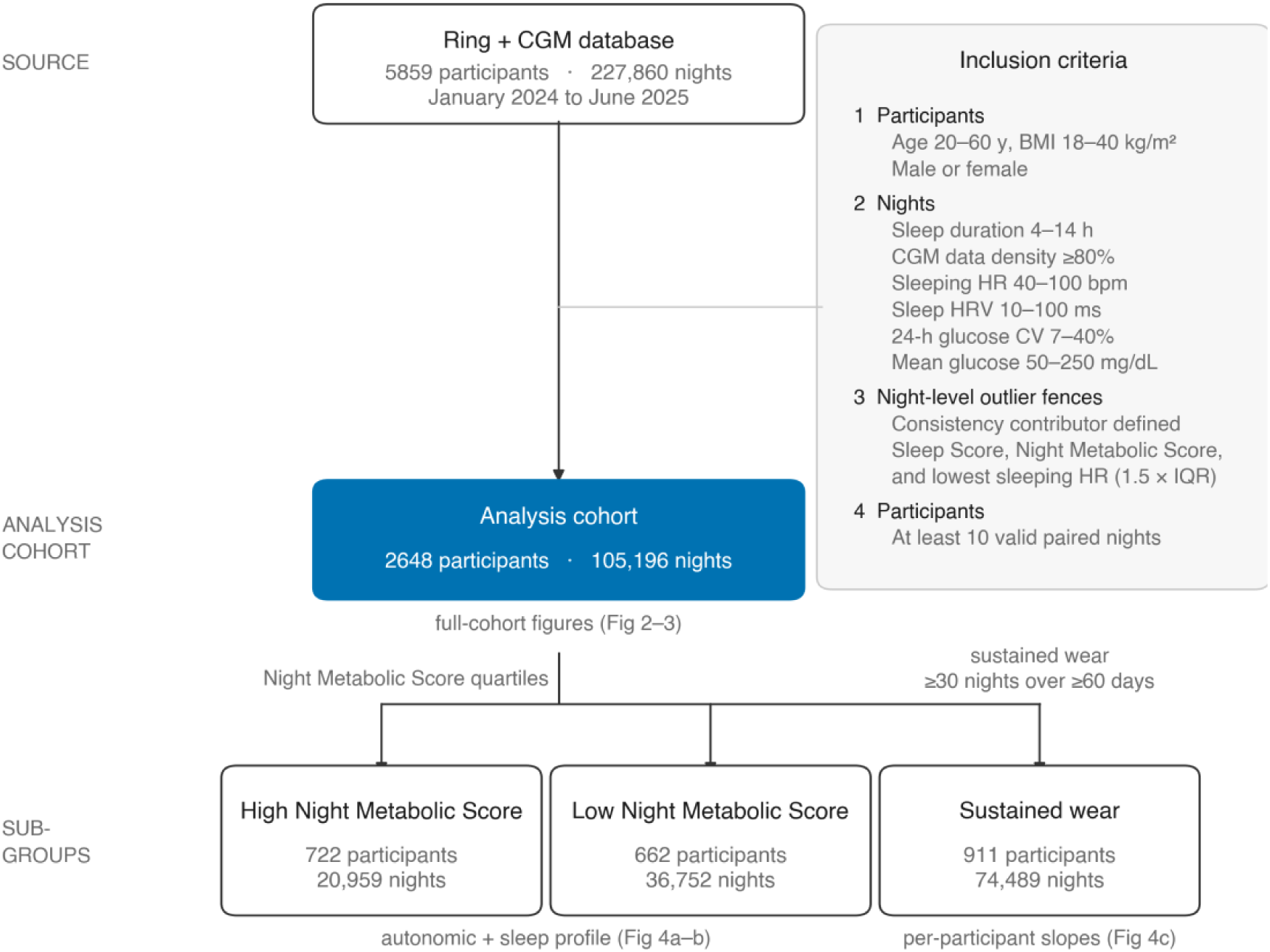
Participant flow from co-monitored ring and CGM records to the analysis cohort and subgroups. Numbers of participants and nights are shown for the source records and the analysis cohort. Inclusion criteria are listed in the order applied (Methods). The lower row shows the subcohorts drawn from the analysis cohort: the metabolic extreme groups and the sustained-wear subcohort. BMI: body mass index; CGM: continuous glucose monitor; CV: coefficient of variation; HR: heart rate; HRV: heart rate variability; IQR: interquartile range.

Nights were further restricted to physiological values typical of nominally healthy adults: average sleeping HR of 40 to 100 bpm [36,37], sleep HRV of 10 to 100 ms [38,39], 24-hour glucose CV of 7% to 40%, and daily and nocturnal mean glucose of 50 to 250 mg/dL. Nights with an undefined consistency contributor were removed, as were nights on which the Sleep Score or Night Metabolic Score lay beyond 1.5 times the interquartile range (IQR), yielding retention ranges of 47.5 to 100 and 38.5 to 100, respectively. The same IQR rule applied to lowest sleeping HR retained nights between 40 and 82 bpm.

Filters were applied in a fixed order: participant-level demographic criteria, night-level sleep and physiological bounds, night-level outlier fences, and finally a participant-level minimum of 10 valid paired nights. The order matters because the participant-level minimum must be evaluated on the nights that survive the night-level filters. The 10-night threshold accommodates the most common 14-day CGM wear period [33] while allowing for real-world gaps in wear.

The glucose bounds, with the participant-level minimum re-evaluated after them, removed 419 nights (0.40%) and 3 participants.

After all filters, the analysis dataset comprised 105,196 nights from 2648 participants; all analyses used this dataset unless stated otherwise.

Two subcohorts were derived. Metabolic extreme groups comprised participants in the top (n=722) and bottom (n=662) quartiles of participant-median Night Metabolic Score (cut-points 94.0 and 80.9; the top group is larger because of ties at the upper cut-point). A sustained-wear subcohort comprised participants with at least 30 nights spanning at least 60 days (n=911), so that per-participant slopes were estimated over a sustained period rather than a single period of wear.

### Statistical Analysis

Analyses were conducted in Python 3.11.8. Mixed-effects models were fitted with statsmodels 0.14.5 [40] using the L-BFGS optimizer, and rank correlations and hypothesis tests with SciPy 1.16.2 [41]. Central tendencies were reported as medians and variability as IQRs, because many metrics were bounded and nonnormal. Effect estimates were reported with 95% confidence intervals. Statistical significance referred to a two-sided threshold of *P*<.05. Reporting follows the STROBE (Strengthening the Reporting of Observational Studies in Epidemiology) guideline for observational cohort studies (Multimedia Appendix 2).

#### Within-Participant and Between-Participant Decomposition

Because each participant contributed multiple nights, tests treating nights as independent observations would be pseudoreplicated. All primary analyses therefore used linear mixed-effects models with a random intercept for participant.

Each predictor was decomposed by person-mean centering into a between-participant component (the participant’s mean across nights) and a within-participant component (the night’s deviation from that mean). Both components were entered simultaneously, with fixed-effect adjustment for age, BMI, and sex. In Wilkinson notation, the model was

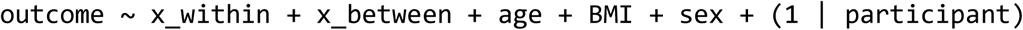

where x_between is the participant’s mean of the predictor and x_within the night’s deviation from it. Continuous covariates were standardized so that they did not dominate the model scale, and predictors were scaled to interpretable increments: per 10 bpm for HR, per 10 ms for HRV, and per 30 minutes for sleep irregularity. The model was also refitted within strata of BMI (18 to <25, 25 to <30, and 30 to 40 kg/m²), age (20-29, 30-39, 40-49, and 50-60 years), and sex, and sex differences were tested with interaction terms between sex and each component.

The between-participant slope estimates whether participants with higher habitual values of the predictor have higher outcome values, corresponding to a conventional cross-sectional comparison. The within-participant slope estimates whether nights on which a participant’s predictor is higher than usual are accompanied by higher-than-usual outcome values, and is derived only from that participant’s night-to-night variation. It is therefore not confounded by time-invariant participant characteristics, such as age, sex, or habitual fitness, but remains susceptible to confounding by factors that vary from night to night. A within-participant slope of the same sign as the between-participant slope indicates that the association is not attributable solely to stable between-person differences.

#### Night-to-Night Sleep Irregularity

Sleep irregularity was defined for each night, as the absolute deviation of that night’s value from the participant’s own median, for sleep duration, sleep onset, and wake time. Unlike a per-participant standard deviation (SD), which is a single value per participant, a night-level deviation can be decomposed into within-and between-participant components. The between-participant component indexes how far a participant’s nights typically depart from their habitual pattern (the conventional irregular-sleeper trait), and the within-participant component indexes a night that departs unusually far from that pattern.

Because sleep onset and wake times were recorded in Coordinated Universal Time for the entire cohort spanning 87 countries, clock-time irregularity was computed circularly: each participant’s center was taken as the circular mean of their clock times corrected by the median of the wrapped deviations, and each night’s irregularity as the circular distance from that center, bounded by 12 hours. A fixed-hour wrap anchor was not used because in a globally distributed cohort any fixed hour falls within some participants’ habitual sleep-onset distribution and generates spurious deviations of nearly 24 hours. Duration irregularity, which is not circular, was the absolute deviation from the participant’s own median. Each irregularity metric was decomposed with the same mixed model.

#### Geographic Region

Because the ring has no GPS, each participant was assigned to the country in which they recorded the largest number of days, using the time zone of the paired smartphone stored with each day’s data. Time zones were mapped to ISO 3166 countries with the public IANA time zone database. Participants spent a median of 98% of their days in their assigned country.

Countries were grouped using the World Bank regional classification, an external scheme adopted to avoid data-driven grouping. Two World Bank regions were split so that stratum labels described their participants: Central Asia was separated from Europe and Central Asia, and North Africa from the Middle East and North Africa. Participants in the separated subregions were retained in the pooled stratum described below, so the cohort remained 2648 participants.

Four regions had at least 100 participants, below which the between-participant slope was too imprecise to compare, and are reported separately: North America, Europe, South Asia, and the Middle East. Three were dominated by one country: the United States made up 98% of North America, India 99.6% of South Asia, and the United Arab Emirates 82% of the Middle East. The remaining 150 participants (East Asia and Pacific, Sub-Saharan Africa, Latin America and Caribbean, North Africa, and Central Asia) were pooled into a single stratum, Other regions, so that no participant was excluded from the comparison. No stratum with fewer than 40 participants was fitted except the Middle East upper glucose tertile (n=23), which is shown as provisional in Figure S2g in Multimedia Appendix 1 and was excluded from heterogeneity tests. Regional composition is given in Table S2 in Multimedia Appendix 1.

The primary model was refitted separately within each region, rather than fitted once with region interaction terms, so that residual and between-participant variances could differ by region. Consistency across the 5 disjoint regional strata was summarized with Cochran Q and I² [42]. Because raw-unit slopes scale with the night-to-night variability of the outcome, which differed by region, the regional models were also refitted on a standardized scale: each within-participant slope was rescaled by the within-participant SDs of glucose and HR in that stratum, expressing it as SDs of a participant’s own night-to-night glucose per SD of their own night-to-night HR. To separate region from glycemic case-mix, the standardized model was further refitted within tertiles of participant mean nocturnal glucose for the 4 named regions. The tertile cut-points (91.0 and 103.6 mg/dL) divided nights rather than participants into thirds, so the tertiles held 1176, 861, and 611 participants. Two sensitivity analyses were performed. The first restricted the sample to participants who spent at least 80% of their days in their assigned country, to assess the effect of travel on regional assignment. The second assessed Ramadan as a potential confounder, because overnight eating patterns change during Ramadan in some regions; models were refitted after removing nights that fell within Ramadan, for the pooled cohort and for each region. No night was excluded from the primary analysis on this basis.

Because regional strata comprise self-selected device users rather than population samples, measured 24-hour mean glucose in each country contributing at least 40 participants was compared by rank with age-standardized diabetes prevalence for 2022 from the World Health Organization Global Health Observatory [43], derived from pooled population-representative surveys [44], and by level with published CGM reference cohorts.

Regional stratification was applied to the HR and sleep-irregularity analyses only. It was not applied to the metabolic extreme groups, which are defined by the outcome and would yield cells too small to interpret.

#### Metabolic Extreme Groups and Per-Participant Coupling

For the extreme-group comparison, each metric was aggregated per participant, using the median for level metrics and, for variability metrics, the mean deviation from the participant’s own center. Subsequently the metrics were z-scored to the full cohort so that heterogeneous metrics shared a common scale. Group means are shown with 95% CI and compared with the Mann-Whitney U test at the participant level, so that each participant contributed one observation.

For the per-participant coupling, an ordinary least-squares slope of nocturnal mean glucose on average sleeping HR was fitted separately within each sustained-wear participant. The sustained-wear definition ensured that every slope was estimated from at least 30 paired nights with nonzero HR variance, so no additional threshold was imposed. The distribution of slopes is summarized by its median, IQR, and 5th to 95th percentiles and was tested against zero with the Wilcoxon signed-rank test.

#### Robustness

A high-leverage observation is one with an extreme predictor value, which can pull a fitted slope disproportionately toward itself. Because a small number of such observations could drive the regression slopes, each primary slope was re-estimated under a series of specifications. Because leverage depends on the predictor, sensitivity to it was assessed by trimming the predictor distribution: dense-core refits restricted to the 10th to 90th and 5th to 95th percentiles of HR, with within-participant deviations additionally capped at 5, 3, and 2 bpm and restricted to the middle 8 deciles. An association driven by extreme predictor values would be expected to attenuate toward zero under trimming.

Additional specifications comprised winsorization at the 2.5th and 97.5th percentiles; removal of the 1% most influential observations by Cook distance; Huber and Theil-Sen robust regression, which assess sensitivity to the outcome tail rather than to leverage; and a participant-cluster bootstrap (1000 resamples) for CIs that respect clustering by participant.

Each specification was fitted to the same 2 components as the primary model: participant means adjusted for age, BMI, and sex, and nightly deviations from each participant’s own mean. Attenuation under a robust estimator was measured against the least-squares fit to the same component.

#### Multiplicity and Joint Testing

The 3 nocturnal glucose outcomes (mean glucose, CV, and time in range) are correlated summaries of the same nightly glucose distribution rather than independent end points and were therefore tested jointly as well as separately. A multivariate analysis of variance of the outcome vector on average sleeping HR provided a single omnibus test, and Spearman correlations among the outcomes are reported to quantify redundancy. Each outcome was also re-estimated with adjustment for the other 2 outcomes to quantify its unique contribution, using least-squares fits without covariates.

## Results

### Participant Characteristics

The analysis cohort comprised 2648 participants from 87 countries contributing 105,196 nights of paired ring and CGM data, a median of 22 (IQR 13-44) nights per participant; 1975 (74.6%) were male (Table 1).

Nightly physiological measures were within ranges reported for healthy adults (Table 1), including average and lowest sleeping HR [45,46] and sleep HRV [47], and median scored sleep duration (7.0 hours) was at the lower bound of the recommended 7 to 9 hours [48]. Nocturnal mean glucose was within the range reported for nondiabetic individuals [49], and median nocturnal glucose CV (8.9%) was lower than the 24-hour CV of 17% (SD 3%) reported for screened healthy individuals [49], consistent with the absence of meals and physical activity during the sleep window. Median nocturnal time in range was 82% (IQR 42%-100%) over the narrower 70 to 110 mg/dL range (Methods).

Values are median (IQR) unless shown as n (%). Demographics are at the participant level; sleep and glucose measures are per night. Sleep duration is scored sleep, which excludes wake after sleep onset and is shorter than the sleep-onset-to-offset interval (Table S3 in Multimedia Appendix 1). Night and Daily Metabolic Score are the platform’s overnight and 24-hour indices, computed from CGM data. BMI: body mass index; CV: coefficient of variation; HR: heart rate; HRV: heart rate variability.

### Sleeping Heart Rate and Nocturnal Glucose

Higher average sleeping HR was associated with higher nocturnal mean glucose at both levels of the decomposition (Figure 2a). Each 10-bpm increment was associated with a 5.7 mg/dL (95% CI 5.4-5.9) higher nocturnal mean glucose within participants and a 6.5 mg/dL (95% CI 5.6-7.3) higher value between participants. Nocturnal glucose CV increased (within participants: +1.37 percentage points, 95% CI 1.30-1.43; between participants: +0.83 percentage points, 95% CI 0.70-0.95; Figure 2b), and nocturnal time in range decreased (within: −6.3 percentage points, 95% CI −6.7 to −6.0; between: −8.2 percentage points, 95% CI −9.2 to −7.2; Figure 2c). All 6 slopes were significant (*P*<.001). A 10-bpm increment corresponded to 2.24 within-participant SDs of sleeping HR; a 1-SD within-participant increase was therefore associated with approximately 2.5 mg/dL higher nocturnal mean glucose. Decile-binned between-participant associations and within-participant night densities are shown in Figure S3 in Multimedia Appendix 1.

**Figure 2.**
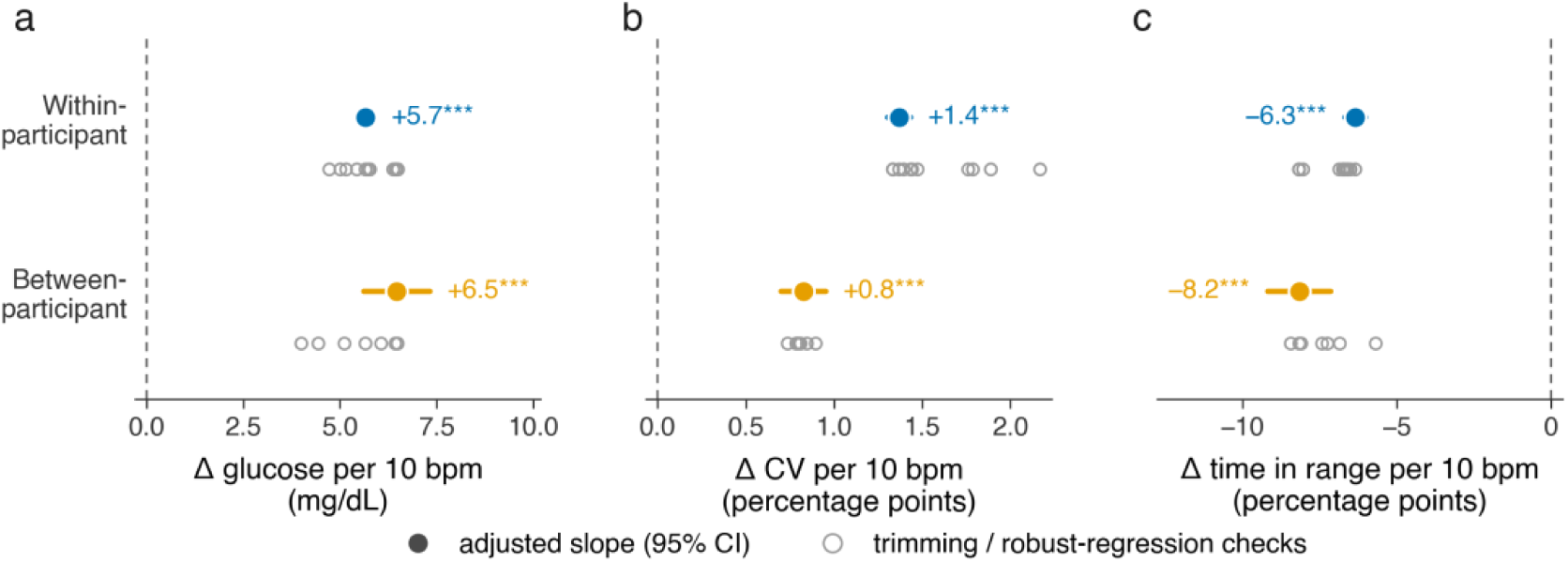
Within-and between-participant associations of average sleeping heart rate with three nocturnal glucose outcomes. One panel per outcome: nocturnal mean glucose **(a)**, nocturnal glucose coefficient of variation **(b)**, and nocturnal time in range **(c)**. Higher average sleeping heart rate was associated with higher mean glucose and coefficient of variation and lower time in range, both within and between participants. Within- and between-participant slopes are from the person-mean-centered mixed model adjusted for age, body mass index, and sex (Methods), per 10 bpm. Filled points are the adjusted slope and whiskers the 95% confidence interval; the dashed line marks a null slope of zero; asterisks give the mixed-model significance of each slope (*** *P*<.001). Gray rings are the same slope under each trimming and robust-regression specification (Methods), shown to indicate the spread across specifications (n=2648 participants, 105,196 nights). CV: coefficient of variation.

Across all trimming and robust-regression specifications, every primary slope retained its sign and its 95% CI excluded the null (Figure 2, gray rings; Multimedia Appendix 1). Trimming the tails of the HR distribution left the within-participant slopes unchanged or steeper and moved the between-participant slopes by no more than 12%. Estimators robust to the outcome tail (Huber and Theil-Sen regression) reduced the between-participant slopes by 3% to 38% and changed the within-participant slopes by at most 17%. Participant-cluster bootstrap 95% CIs for nocturnal mean glucose were 4.9-6.4 mg/dL (within participants) and 5.5-7.3 mg/dL (between participants) per 10 bpm.

The within-participant estimate was insensitive to adjustment for time-invariant covariates. Across 5 specifications of age, from omission to a 4-level factor, and with or without adjustment for sex, the within-participant slope was +5.661 mg/dL per 10 bpm in every model (Table S4 in Multimedia Appendix 1). The association was present in every BMI stratum (normal weight, +4.8; overweight, +5.8; obese, +6.5 mg/dL per 10 bpm) and every age stratum (20-29 years, +5.3; 30-39 years, +5.1; 40-49 years, +5.6; 50-60 years, +6.4). Sex was a negative confounder of the between-participant estimate: omitting it reduced the slope from +6.47 to +5.73 mg/dL per 10 bpm.

Sex also modified the association. The within-participant slope was +6.2 mg/dL per 10 bpm in men (n=1975) and +3.3 in women (n=673; sex interaction *P*<.001 at both levels), a 1.9-fold difference that was attenuated to 1.6-fold, but not eliminated, by rescaling with each sex’s within-participant SDs of glucose and HR, and that widened within the normal-weight stratum (Table S4 in Multimedia Appendix 1).

### Redundancy Among the Glucose Outcomes

Across participant medians, time in range correlated with mean glucose (Spearman ρ=-0.49) and CV (ρ=-0.61), and mean glucose correlated weakly with CV (ρ=0.22); the corresponding within-participant coefficients were −0.41, −0.29, and 0.05. A multivariate test of the outcome vector on average sleeping HR yielded a single omnibus between-participant association (Wilks λ=0.89, F_3,2641_=106.0, *P*<.001; canonical R=0.33).

Under mutual adjustment for the other 2 outcomes, all between-participant associations attenuated toward the null, consistent with a single shared glycemic axis (Figure S4 in Multimedia Appendix 1). Within participants, the associations with mean glucose and CV were largely retained (from 0.35 to 0.27 and from 0.29 to 0.26 outcome SDs per 10 bpm), whereas the association with time in range was largely absorbed (from −0.26 to −0.05). Because the outcomes are co-measurements of one distribution, mutual adjustment indexed redundancy rather than confounding.

### Alternative Autonomic Metrics

Lowest sleeping HR showed the same pattern as average sleeping HR (within participants: +3.5 mg/dL per 10 bpm, 95% CI 3.2-3.7; between participants: +7.5, 95% CI 6.4-8.5). Sleep HRV was inversely associated with nocturnal mean glucose (within: −1.5 mg/dL per 10 ms, 95% CI −1.7 to −1.4; between: −2.4, 95% CI −3.0 to −1.7; Figure S5 in Multimedia Appendix 1). Higher average sleeping HR was associated with a lower Night Metabolic Score (within: − 3.9 points per 10 bpm; between: −4.1), and higher HRV with a higher score (+1.1 and +1.4 points per 10 ms; Figure S6 in Multimedia Appendix 1).

### Night-to-Night Sleep Irregularity

All 3 irregularity metrics showed between-participant associations with nocturnal glycemia, differing in magnitude and in the number of outcomes involved (Figure 3). Sleep-duration irregularity showed the largest associations and was associated with all 3 outcomes: per 30 minutes of irregularity, nocturnal mean glucose was 4.1 mg/dL higher (95% CI 2.6-5.7), CV 0.52 percentage points higher (95% CI 0.30-0.75), and time in range 4.0 percentage points lower (95% CI 2.2-5.8). Wake-time irregularity was associated with all 3 outcomes at smaller magnitudes (mean glucose +1.1 mg/dL, 95% CI 0.4-1.8; CV +0.14 percentage points, 95% CI 0.03-0.24; time in range −1.6 percentage points, 95% CI −2.5 to − 0.8). Sleep-onset irregularity was associated with CV (+0.12 percentage points, 95% CI 0.01-0.22) and time in range (−1.1 percentage points, 95% CI −2.0 to −0.2) but not with mean glucose (+0.67 mg/dL, 95% CI −0.06 to 1.40).

**Figure 3.**
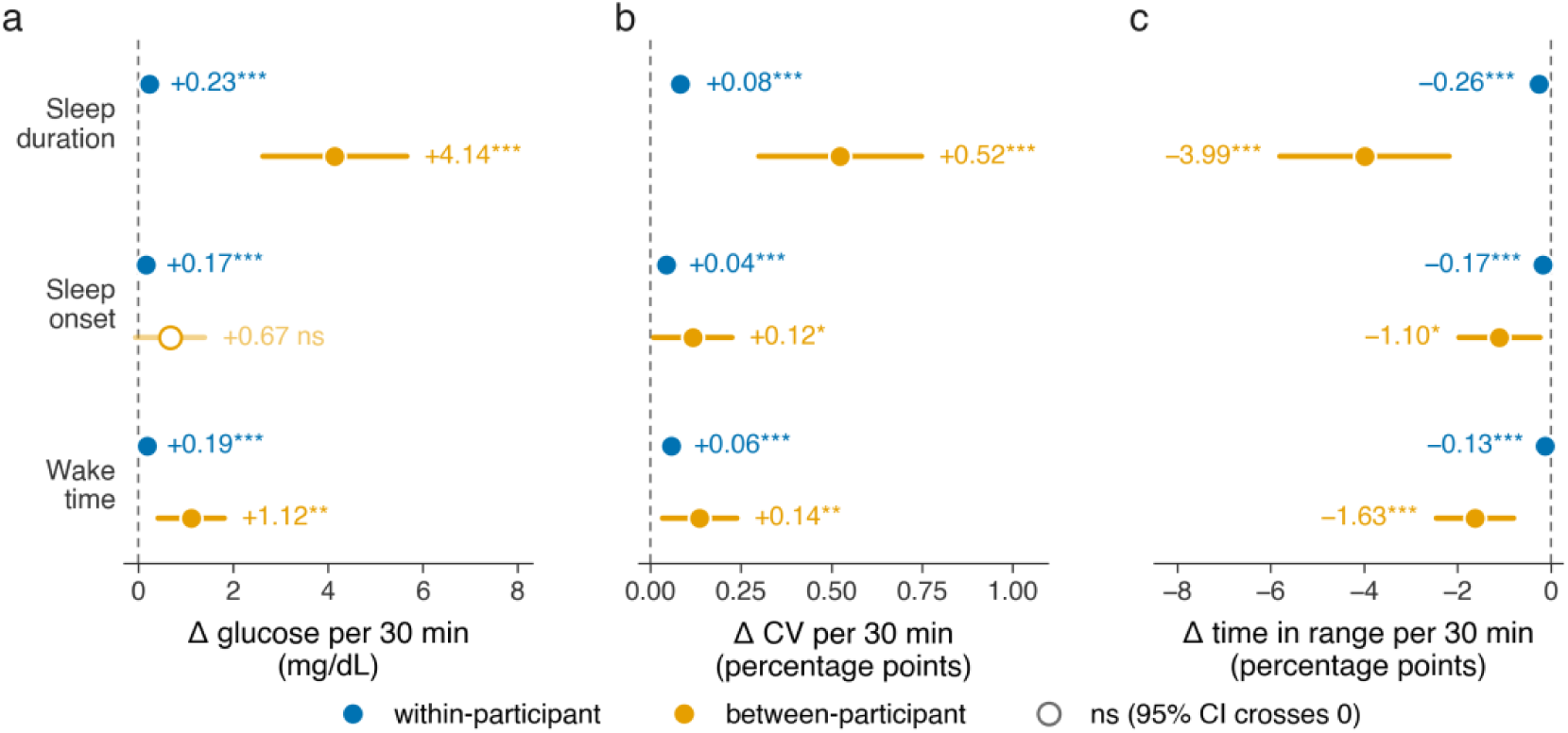
Within- and between-participant associations of night-to-night sleep irregularity with three nocturnal glucose outcomes. One panel per outcome: mean glucose **(a)**, glucose coefficient of variation **(b)**, and time in range **(c)**. Sleep-duration irregularity was associated with all 3 outcomes between participants, and the within-participant slopes were small. Irregularity is each night’s deviation from the participant’s own center for scored sleep duration, sleep onset, and wake time, computed circularly for the 2 clock times (Methods). Slopes are per 30 minutes of irregularity, decomposed and drawn as in Figure 2; open markers denote a slope whose interval crosses the null; asterisks give mixed-model significance (*** *P*<.001, ** *P*<.01, * *P*<.05; ns, not significant) (n=2648 participants, 105,196 nights). CV: coefficient of variation.

Within-participant associations with nocturnal mean glucose were significant for all 3 metrics but small (duration, +0.23; wake time, +0.19; onset, +0.17 mg/dL per 30 minutes), 4- to 18-fold smaller than the corresponding between-participant slopes and smaller than the HR associations (Figure 2).

The irregularity metrics were correlated but not redundant. Between participants the 3 measures correlated moderately with one another (ρ=0.54 to 0.63), and within participants weakly (ρ=0.19 to 0.28; Figure S7 in Multimedia Appendix 1). Under mutual adjustment between participants, duration irregularity remained associated with all 3 outcomes, wake-time irregularity only with time in range, and onset irregularity with none (Figure S8 in Multimedia Appendix 1). The Sleep Score correlated weakly with the Night Metabolic Score between participants (ρ=0.23; Figure S9 in Multimedia Appendix 1).

### Consistency Across Geographic Regions

The association between average sleeping HR and nocturnal glucose had the same direction, with 95% CIs excluding the null, in all 5 regional strata for all 3 outcomes at both levels (29/30 estimates *P*<.001; 1/30 *P*<.01; Figure S2 in Multimedia Appendix 1). Within-participant slopes for nocturnal mean glucose were +5.05 mg/dL per 10 bpm in North America (n=796), +4.29 in Europe (n=783), and +8.64 in South Asia (n=710); they were +2.14 in the Middle East (n=209) and +5.22 in Other regions (n=150). Magnitude was heterogeneous across regions within participants (mean glucose: Cochran Q_4_=329.1, I²=99%) and, to a lesser extent, between participants (Q_4_=29.5, I²=86%). On the standardized scale (Methods), the range of regional estimates narrowed from 4.0-fold to 2.8-fold (Multimedia Appendix 1). Restriction to the 2009 participants who spent at least 80% of their days in their assigned country changed 4 out of 5 regional estimates by 0.4 mg/dL per 10 bpm or less (Middle East: +2.1 to +4.0), and exclusion of 12,408 nights falling within Ramadan left the pooled within-participant slope unchanged (+5.7 mg/dL per 10 bpm; Multimedia Appendix 1).

The within-participant slope increased across tertiles of participants’ mean nocturnal glucose, from +2.5 mg/dL per 10 bpm in the lowest to +9.4 mg/dL per 10 bpm in the highest tertile. On the standardized scale, the gradient was smaller but persisted (+0.107, +0.155, and +0.189 SD per SD), and differences among the 4 named regions remained significant within every tertile (all *P*<.001; Figure S2g in Multimedia Appendix 1).

The between-participant association of sleep-duration irregularity with nocturnal glycemia was directionally consistent across the 4 primary regions for all 3 outcomes. Its 95% CIs excluded the null in North America and South Asia but not consistently in Europe or the Middle East (Table S5 in Multimedia Appendix 1).

Measured 24-hour mean glucose was weakly rank-correlated with national age-standardized diabetes prevalence across the 8 countries contributing at least 40 participants (Spearman ρ=0.40, *P*=.32). The United Arab Emirates had the highest national prevalence but the third-lowest measured glucose, and its exclusion increased the correlation to 0.71 (Table S6 in Multimedia Appendix 1). The cohort’s 24-hour mean glucose (103.8 mg/dL) lay between published CGM reference values for screened healthy individuals (98.5 and 102.4 mg/dL) and adults with prediabetes (112.2 mg/dL) [35,49].

### Metabolic Extreme Groups and Per-Participant Coupling

Compared with the low Night Metabolic Score group (n=662), the high-score group (n=722) had lower average and lowest sleeping HR, higher sleep HRV, longer sleep duration, higher Sleep Scores, and lower night-to-night irregularity in sleep duration, sleep onset, and wake time (Figures 4a and 4b; all *P*<.001). The groups differed by 36.7 mg/dL in participant-median nocturnal glucose and by 7.0 bpm in average sleeping HR. Because the groups were defined on nocturnal glycemia, this comparison was descriptive.

**Figure 4.**
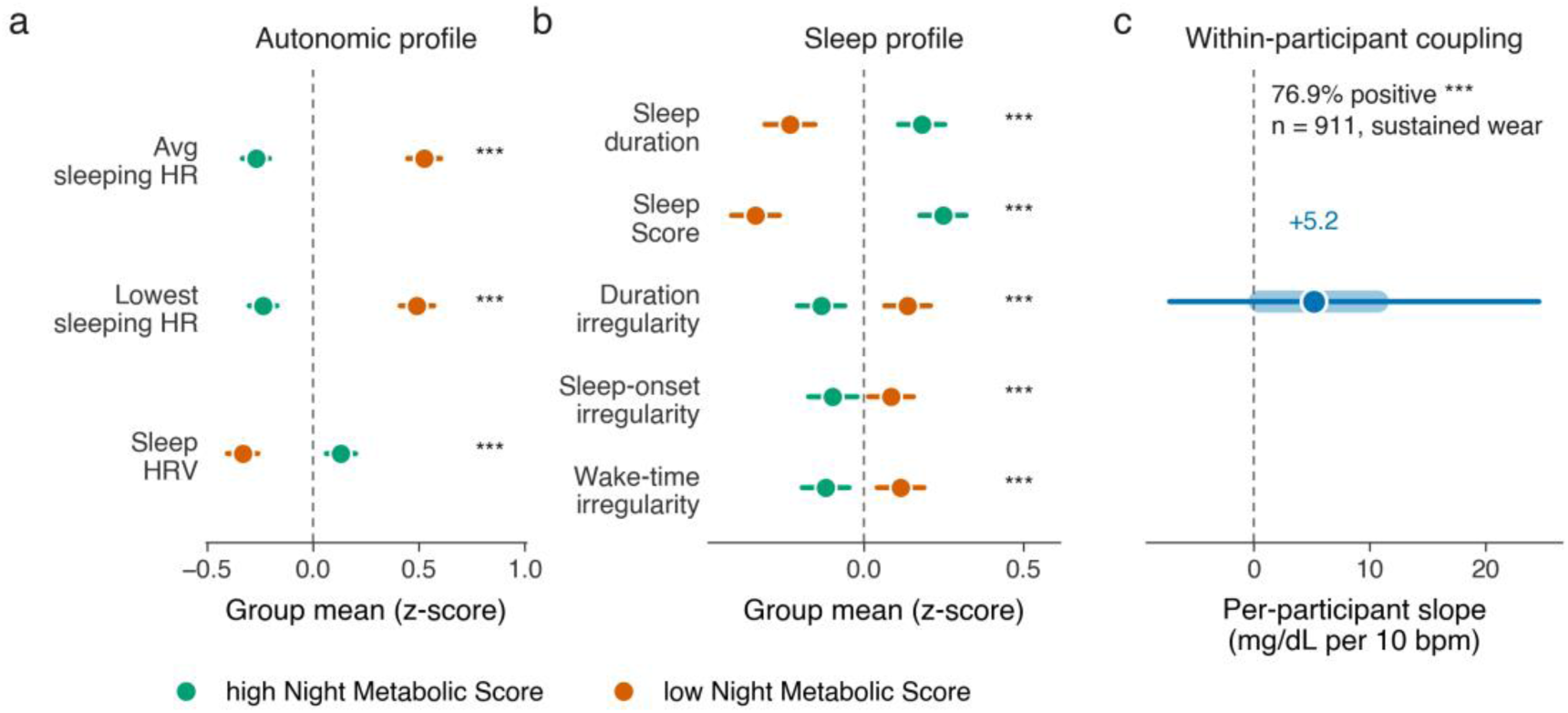
Profiles of the metabolic extreme groups and the distribution of per-participant coupling. **(a)** Autonomic and **(b)** sleep profiles of participants in the top (high) and bottom (low) quartile of median Night Metabolic Score, expressed as standardized group means (z-scores relative to the full cohort) with 95% confidence intervals. Metrics are aggregated per participant as described in Methods. The high group had lower sleeping heart rate, higher heart rate variability, and more regular sleep, and most sustained-wear participants had a positive slope. Asterisks to the right of each metric give the high-versus-low group difference (Mann-Whitney; *** *P*<.001). **(c)** Per-participant ordinary least-squares slopes of nocturnal mean glucose on average sleeping heart rate in sustained-wear participants: median (point), interquartile range (thick bar), and 5th to 95th percentiles (thin line), with the dashed line at zero (high, 722 participants; low, 662; sustained-wear, 911). Avg: average; HR: heart rate; HRV: heart rate variability.

Among the 911 sustained-wear participants (median 60 nights each), the median per-participant slope of nocturnal mean glucose on average sleeping HR was +5.2 mg/dL per 10 bpm (IQR 0.5-10.7), and 701 of 911 participants (76.9%, 95% CI 74.1%-79.6%) had a positive slope (Wilcoxon signed-rank *P*<.001; Figure 4c). The sustained-wear subcohort was comparable with the full cohort in median age (44 vs 42 years), BMI (26.8 vs 26.3 kg/m²), proportion of men (78.0% vs 74.6%), average sleeping HR (within 1 bpm), and nocturnal mean glucose (within 2 mg/dL).

## Discussion

In this free-living cohort of 2648 adults, higher average sleeping HR was associated with higher nocturnal mean glucose, higher nocturnal glucose variability, and lower time in range. Each association was present both between and within participants. The within-participant estimate (+5.7 mg/dL per 10 bpm) was of similar magnitude to the between-participant estimate (+6.5 mg/dL per 10 bpm). Because it is derived only from each participant’s night-to-night variation, it cannot be attributed to time-invariant characteristics such as adiposity, age, or sex. The association was positive in 76.9% of sustained-wear participants and directionally consistent across all BMI, age, and regional strata. Its magnitude at typical night-to-night variation was modest, approximately 2.5 mg/dL per within-participant SD of sleeping HR. In contrast, night-to-night irregularity in sleep duration was associated with nocturnal glycemia predominantly between participants.

The between-participant association is consistent with reports linking higher resting HR to lower insulin sensitivity in nondiabetic adults [7], to blood glucose in relation to cardiac autonomic function [8], and to subsequent changes in glucose metabolism [9]; the present study adds an estimate of magnitude from 105,196 free-living nights. The inverse association with sleep HRV agrees with modest inverse HRV-glucose associations during sleep in small laboratory cohorts [2] and with associations between sleep-stage HRV and glycemic control in type 2 diabetes [4]. These studies estimated associations across individuals and could not distinguish a stable trait from a night-varying state. Laboratory studies have described within-individual temporal coupling between sleep microarchitecture, autonomic state transitions, and interstitial glucose [21,22]. The present findings are aligned with that work at whole-night resolution but do not test the proposed mechanisms, because no within-night features or hormonal intermediates were analyzed. Testing them would require finer-grained analysis alongside blood-based biomarkers.

Two features of the design extend this literature. First, the density of paired nights (median 22 per participant, and 911 sustained-wear participants) allowed within-and between-participant components to be estimated jointly and per-participant slopes to be characterized, which is not feasible in smaller cohorts [2,21–23]. Second, the cohort spanned 87 countries, permitting a test of regional consistency not reported in those studies.

Interpretation of the within-participant estimate is constrained by the design. Person-mean centering removes time-invariant confounding but not confounding by factors that vary from night to night, including alcohol intake, evening meal timing and composition, acute illness, psychological stress, medication changes, and undiagnosed sleep-disordered breathing. None of these was available in the study data. HR and glucose were measured concurrently over the same night, so no temporal ordering was established. The within-participant association therefore indicates that sleeping HR and nocturnal glucose covary as part of a shared nightly state. It does not establish that autonomic tone influences glucose, and it does not support inferring an individual’s glycemia from a single night’s HR. The between-participant estimate is additionally subject to residual confounding by stable unmeasured characteristics, including chronotype, shift work, and menopausal status.

The coupling was not uniform. It was steeper at higher glucose and differed across regions; both gradients narrowed but persisted on the standardized scale, and glycemic case-mix contributed to the regional differences. The regional strata also represent device users rather than national populations with the measured glycemia corresponding only weakly to national diabetes prevalence. Participants were self-selected consumers able to afford, and motivated to use, both devices, and 74.6% were male. The presence and direction of the association therefore was replicated across strata, whereas its magnitude should not be extrapolated to populations with different glycemic distributions.

Menstrual cycle phase was not available in the study data and may contribute to the weaker within-participant association in women. It varies within participants across the recording window and is associated with changes in both HR and glucose, so unmodeled cyclical variation could attenuate the within-participant estimate. The sex difference is therefore best interpreted as a difference in the measured association rather than as evidence of sex-specific physiology, and studies that record cycle phase are needed to distinguish the two.

Sleep regularity is increasingly regarded as a health target. Irregular sleep duration has been associated with incident type 2 diabetes [50], with greater regularity relating to lower all-cause and cardiometabolic mortality [51]. Sleep health is also included in the American Heart Association’s Life’s Essential 8 [52]. The between-participant association of sleep-duration irregularity with nocturnal glycemia is consistent with this literature, which likewise contrasts habitually irregular with habitually regular sleepers [12,15,17,19,50,51]. Such comparisons do not describe the effect of a single irregular night on an individual, and the corresponding within-participant association here was small (+0.23 vs +4.1 mg/dL per 30 minutes between participants).

Associations of this size reach statistical significance with 105,196 nights. They are consistent with a study of 69 healthy university students followed for up to 14 days, in which night-to-night variation in sleep did not significantly change nocturnal mean glucose, and the association of longer sleep with lower same-night glucose variability was judged too small to be functionally significant [23]. The predictors differ: that study modeled each night’s sleep duration, timing, and efficiency, whereas this study modeled each night’s deviation from the participant’s own median. In this study, each participant’s habitual irregularity was estimated from a median of 22 nights. Sampling error in these estimates is expected to attenuate the between-participant slopes, so the between-within asymmetry reported here is unlikely to be overstated. Sleep-duration irregularity is therefore best regarded as a correlate of nocturnal glycemia; whether modifying it alters glycemia would require prospective manipulation of sleep in free-living settings.

In the descriptive extreme-group comparison, the 7.0-bpm between-group difference in sleeping HR corresponds, at the within-participant slope, to approximately 4.0 mg/dL, or 11% of the 36.7 mg/dL difference in nocturnal glucose. The per-participant slopes, which are independent of group assignment, provide the more appropriate estimate of individual-level coupling.

Several measurement considerations also apply. HR and HRV were derived with proprietary algorithms on a single consumer device. Ring HR agreed with 2 independent references, including an FDA-cleared device (mean absolute error 2.4 bpm; Table S1 and Figure S1 in Multimedia Appendix 1). BMI was self-reported, so the gradient across BMI strata (+4.8 to +6.5 mg/dL per 10 bpm) should be read as a trend rather than a calibrated dose-response relationship. The 70 to 110 mg/dL range used for time in range has been shown to discriminate normoglycemia from prediabetes in a single screened cohort [35]. We do not present the reduced range as a clinically validated measure. Glucose metrics were computed over single nights rather than the standard 14-day window [33,34] and thus carry greater within-person variability. Because this variability is in the outcome, it widens the confidence intervals rather than biasing the slopes, and the intervals remained narrow. Estimators robust to the outcome tail attenuated the between-participant slopes by up to 38%, so participants with the highest nocturnal glucose contribute disproportionately to the pooled estimates. Even so, every primary slope kept its sign and its 95% CI excluded the null under these estimators (Figure 2). Finally, the outlier fences on the Night Metabolic Score, which is computed from the same nocturnal glucose measures that served as outcomes, were wide (38.5-100) and intended to remove implausible nights, and thus unlikely to affect key results reported here.

### Conclusions

Consumer smart rings and CGMs are designed to track an individual’s physiological patterns over time. Across a large, heterogeneous, and global population studied in free-living conditions, nights on which a participant’s average sleeping HR exceeded their own mean were accompanied by higher nocturnal glucose, and this within-participant coupling was directionally consistent across BMI, age, and geographical strata. It is therefore not attributable solely to stable between-person differences and is the class of pattern that repeated nightly measurement is positioned to resolve. Its modest size points to a trend detectable across many nights rather than a signal interpretable from a single night. Sleep-duration irregularity, in contrast, was associated with nocturnal glycemia mainly between participants. These findings are associational and do not establish that modifying sleeping HR or sleep regularity would alter nocturnal glucose.

## Authors’ Contributions

Conceptualization: AB, HPK Data curation: DG

Formal analysis: ND (lead), DG (supporting; ran the analyses for an earlier preprint of this study [53])

Investigation: MD

Methodology: AB, HPK, ND

Resources: MPS, TW

Supervision: AB

Writing – original draft: ND (equal), AB (equal)

Writing – review & editing: ND, DG, HPK, MD, MPS, TW, AB

## Funding

The study was supported by Ultrahuman Healthcare Private Limited internal research funds. Apart from the work of the authors it employed, the funder had no role in the study design, analysis, interpretation, or decision to publish.

## Conflicts of Interest

At the time of data analysis, AB, ND, and DG were employees of Ultrahuman Healthcare Private Limited; HPK was an advisor to Ultrahuman Healthcare Private Limited; MD was retained on a contract for specific research analysis and collaborative activities; MPS and TW did not receive any material or fiduciary support from Ultrahuman for this project.

TW declares no other conflict of interest.

MPS is a cofounder and scientific advisor of Xthera, Exposomics, Filtricine, Fodsel, iollo, InVu Health, January AI, Marble Therapeutics, Mirvie, Next Thought AI, Orange Street Ventures, Personalis, Protos Biologics, Qbio, RTHM, and SensOmics. MPS is a scientific advisor of Abbratech, Applied Cognition, Enovone, Jupiter Therapeutics, M3 Helium, Mitrix, Neuvivo, Onza, Sigil Biosciences, Captify Inc, WndrHLTH, Yuvan Research, Ovul, Erudio, and Lyten. MPS is an investor and scientific advisor of R42 and Swaza. MPS is an investor in Repair Biotechnologies. These positions are not related to this paper.

## Data Availability

Individual-level data are not publicly available because they are commercial user records subject to privacy protections; aggregate, de-identified data can be made available upon reasonable request. The Sleep Score and Metabolic Score platform codes and technical details are proprietary assets of Ultrahuman and will not be disclosed. ND stands guarantor of the veracity of the study data. For inquiries please contact Nihav Dhawale.

## Code Availability

Analysis code, apart from that containing proprietary information, can be made available upon reasonable request. Please contact Nihav Dhawale.

## Supporting information

Supplemental Information STROBE Checklist

## Acknowledgments

The authors thank the Ultrahuman Ring and continuous glucose monitor users who allowed their de-identified data to be used for research. The authors also thank Aditya Patwari, Nishanth Krishnan, and members of the Ultrahuman engineering and data analytics teams for support in assembling the study data, and Neeraj Kumar, Adhit Shet, and Bhuvan Srinivasan for technical support. The authors also thank Dr Sahil Chopra of Empower Sleep for helpful discussions during early ideation.

We used the generative artificial intelligence tools Claude Opus 5 and Claude Opus 5.5 by Anthropic [54] to write analysis and figure code, to cross-check reported values against the analysis outputs, and to draft and edit manuscript text. The authors directed, reviewed, and revised all output and take full responsibility for the content. The interaction transcripts are not provided because they contain proprietary information.

## Abbreviations

BMI: body mass index
CGM: continuous glucose monitor
CV: coefficient of variation
FDA: US Food and Drug Administration
HR: heart rate
HRV: heart rate variability
IQR: interquartile range
SD: standard deviation
STROBE: Strengthening the Reporting of Observational Studies in Epidemiology

## Multimedia Appendix 1

Supplementary information: device validation, Figures S1-S9, supplementary results, and Tables S1-S6.

## Multimedia Appendix 2

STROBE checklist.

## Notes

### Competing Interest Statement

At the time of data analysis and manuscript preparation. Aditi Bhattacharya (A.B.), Nihav Dhawale (N.D.), Dhruv Gandhi (D.G.), were employees of Ultrahuman Healthcare Pvt. Ltd; Hans Peter-Kubis (H.-P.K.) was an advisor to Ultrahuman Healthcare Pvt. Ltd.; Matt Driller (M.D.) was retained on a contract for specific research analysis and collaborative activities; Michael P Snyder (M.P. S.) and Tao Wang (T.W.) did not receive any material or fiduciary support from Ultrahuman for this project.
T.W. declares no other conflict of interest.
M.P.S. is a cofounder and scientific advisor of Xthera, Exposomics, Filtricine, Fodsel, iollo, InVu Health, January AI, Marble Therapeutics, Mirvie, Next Thought AI, Orange Street Ventures, Personalis, Protos Biologics, Qbio, RTHM, SensOmics. M.P.S is a scientific advisor of Abbratech, Applied Cognition, Enovone, Jupiter Therapeutics, M3 Helium, Mitrix, Neuvivo, Onza, Sigil Biosciences, Captify Inc, WndrHLTH, Yuvan Research, Ovul, Erudio and Lyten. MPS is an investor and scientific advisor of R42 and Swaza. MPS is an investor in Repair Biotechnologies. These positions are not related to this paper.

### Author Declarations

This was a real world retrospective observational survey based on data derived from Ultrahuman platform users and adhered to the Ultrahuman terms of use and privacy policy which allows for analysis of de-identified grouped data for scientific research. This can be publicly accessed on the company website. Thus participants consented via the onboarding process on the Ultrahuman platform and continued product use. As the study was non-invasive and involved no dietary sleep or exercise interventions with all reported data de-identified explicit institutional ethics board approval was not required. For participant details for the data reported in Supplementary Figure S1, the study was approved by the La Trobe University Human Ethics Committee number HEC22369.

### Summary of Updates

This version of the manuscript has contains different analysis and associational examination from the previous submission. The main cohort,retrospective analysis approach remains the same.

## References

1. Niethard N, Hallschmid M. A sweet spot for the sleeping brain: linking human sleep physiology and glucoregulation. Cell Rep Med. 2023;4(7):101123. PMID:37467713 doi:10.1016/j.xcrm.2023.101123

2. Kajisa T, Kuroi T, Hara H, Sakai T. Correlation analysis of heart rate variations and glucose fluctuations during sleep. Sleep Med. 2024;113:180–187. PMID:38042028 doi:10.1016/j.sleep.2023.11.038

3. Stamatakis KA, Punjabi NM. Effects of sleep fragmentation on glucose metabolism in normal subjects. Chest. 2010;137(1):95–101. PMID:19542260 doi:10.1378/chest.09-0791

4. Cheng W, Chen H, Tian L, Ma Z, Cui X. Heart rate variability in different sleep stages is associated with metabolic function and glycemic control in type 2 diabetes mellitus. Front Physiol. 2023;14:1157270. PMID:37123273 doi:10.3389/fphys.2023.1157270

5. Knutson KL. Impact of sleep and sleep loss on glucose homeostasis and appetite regulation. Sleep Med Clin. 2007;2(2):187–197. PMID:18516218 doi:10.1016/j.jsmc.2007.03.004

6. Briançon-Marjollet A, Weiszenstein M, Henri M, Thomas A, Godin-Ribuot D, Polak J. The impact of sleep disorders on glucose metabolism: endocrine and molecular mechanisms. Diabetol Metab Syndr. 2015;7:25. PMID:25834642 doi:10.1186/s13098-015-0018-3

7. Festa A, D’Agostino R Jr, Hales CN, Mykkänen L, Haffner SM. Heart rate in relation to insulin sensitivity and insulin secretion in nondiabetic subjects. Diabetes Care. 2000;23(5):624–628. PMID:10834420 doi:10.2337/diacare.23.5.624

8. Valensi P, Extramiana F, Lange C, et al. Influence of blood glucose on heart rate and cardiac autonomic function. The DESIR study. Diabet Med. 2011;28(4):440–449. PMID:21204961 doi:10.1111/j.1464-5491.2010.03222.x

9. Hansen CS, Færch K, Jørgensen ME, et al. Heart rate, autonomic function, and future changes in glucose metabolism in individuals without diabetes: the Whitehall II cohort study. Diabetes Care. 2019;42(5):867–874. PMID:30940642 doi:10.2337/dc18-1838

10. Ford ES, Cunningham TJ, Croft JB. Trends in self-reported sleep duration among US adults from 1985 to 2012. Sleep. 2015;38(5):829–832. PMID:25669182 doi:10.5665/sleep.4684

11. Coutrot A, Lazar AS, Richards M, et al. Reported sleep duration reveals segmentation of the adult life-course into three phases. Nat Commun. 2022;13(1):7697. PMID:36509747 doi:10.1038/s41467-022-34624-8

12. Wu Y, Zhai L, Zhang D. Sleep duration and obesity among adults: a meta-analysis of prospective studies. Sleep Med. 2014;15(12):1456–1462. PMID:25450058 doi:10.1016/j.sleep.2014.07.018

13. ResMed. ResMed’s 2025 global sleep survey. 2025. https://document.resmed.com/documents/global/resmed-wsd-2025-report.pdf [accessed Oct 13, 2026]

14. Kitchlew R, Haider M, Batool S, Farooq F, Shamim Ul Husnain M. Quality of sleep and its associated factors among diabetics and non diabetics. J Ayub Med Coll Abbottabad. 2020;32(4):507–511. PMID:33225653

15. Cappuccio FP, D’Elia L, Strazzullo P, Miller MA. Quantity and quality of sleep and incidence of type 2 diabetes: a systematic review and meta-analysis. Diabetes Care. 2010;33(2):414–420. PMID:19910503 doi:10.2337/dc09-1124

16. van Dijk M, Donga E, van Dijk JG, et al. Disturbed subjective sleep characteristics in adult patients with long-standing type 1 diabetes mellitus. Diabetologia. 2011;54(8):1967–1976. PMID:21573904 doi:10.1007/s00125-011-2184-7

17. Itani O, Jike M, Watanabe N, Kaneita Y. Short sleep duration and health outcomes: a systematic review, meta-analysis, and meta-regression. Sleep Med. 2017;32:246–256. PMID:27743803 doi:10.1016/j.sleep.2016.08.006

18. Wang B, Zhang H, Sun Y, et al. Association of sleep patterns and cardiovascular disease risk is modified by glucose tolerance status. Diabetes Metab Res Rev. 2023;39(6):e3642. PMID:37009685 doi:10.1002/dmrr.3642

19. Jike M, Itani O, Watanabe N, Buysse DJ, Kaneita Y. Long sleep duration and health outcomes: a systematic review, meta-analysis and meta-regression. Sleep Med Rev. 2018;39:25–36. PMID:28890167 doi:10.1016/j.smrv.2017.06.011

20. Byun JI, Cha KS, Jun JE, et al. Dynamic changes in nocturnal blood glucose levels are associated with sleep-related features in patients with obstructive sleep apnea. Sci Rep. 2020;10(1):17877. PMID:33087786 doi:10.1038/s41598-020-74908-x

21. Yang X, Fedumenti FT, Niethard N, Hallschmid M, Born J, Rauss K. Regulation of peripheral glucose levels during human sleep. Sleep. 2025;48(6):zsaf042. PMID:39987458 doi:10.1093/sleep/zsaf042

22. Vallat R, Shah VD, Walker MP. Coordinated human sleeping brainwaves map peripheral body glucose homeostasis. Cell Rep Med. 2023;4(7):101100. PMID:37421946 doi:10.1016/j.xcrm.2023.101100

23. Ng ASC, Tai ES, Chee MWL. Effects of night-to-night variations in objectively measured sleep on blood glucose in healthy university students. Sleep. 2025;48(2):zsae224. PMID:39325824 doi:10.1093/sleep/zsae224

24. Hyun U, Sohn JW. Autonomic control of energy balance and glucose homeostasis. Exp Mol Med. 2022;54(4):370–376. PMID:35474336 doi:10.1038/s12276-021-00705-9

25. Lin EE, Scott-Solomon E, Kuruvilla R. Peripheral innervation in the regulation of glucose homeostasis. Trends Neurosci. 2021;44(3):189–202. PMID:33229051 doi:10.1016/j.tins.2020.10.015

26. Pocai A, Obici S, Schwartz GJ, Rossetti L. A brain-liver circuit regulates glucose homeostasis. Cell Metab. 2005;1(1):53–61. PMID:16054044 doi:10.1016/j.cmet.2004.11.001

27. Abbott. FreeStyle Libre CGM systems for healthcare providers. https://www.freestyleprovider.abbott/us-en/home.html [accessed Oct 13, 2026]

28. Krishnan N, et al. Sleep heart rate sensing by Ultrahuman Ring AIR. Ultrahuman Science Portal. 2026. https://science.ultrahuman.com/studies/sleep-heart-rate-sensing

29. Ultrahuman. Terms of use. https://ultrahumanapp.notion.site/TERMS-OF-USE-7a541a34b77b4cb79566a327578873bb [accessed Oct 13, 2026]

30. Ultrahuman. Privacy policy. https://ultrahumanapp.notion.site/PRIVACY-POLICY-16dccc2b1b2b4c4bb9a462c155b3bef1 [accessed Oct 13, 2026]

31. Driller M, Bodner ME, Fenuta A, Stevenson S, Suppiah H. How does the Sleep Regularity Questionnaire relate to wearable-and diary-derived sleep regularity? J Circadian Rhythms. 2026;24:2. PMID:42145918 doi:10.5334/jcr.265

32. Ehlert B, Aron D, Perelman D, Wu Y, Snyder MP. Glucose360: an open-source Python platform with event-based integration for continuous glucose monitoring data analysis. Diabetes Technol Ther. 2026;28(2):130–141. PMID:40900178 doi:10.1177/15209156251374711

33. Camerlingo N, Vettoretti M, Sparacino G, et al. Design of clinical trials to assess diabetes treatment: minimum duration of continuous glucose monitoring data to estimate time-in-ranges with the desired precision. Diabetes Obes Metab. 2021;23(11):2446–2454. PMID:34212483 doi:10.1111/dom.14483

34. Danne T, Nimri R, Battelino T, et al. International consensus on use of continuous glucose monitoring. Diabetes Care. 2017;40(12):1631–1640. PMID:29162583 doi:10.2337/dc17-1600

35. Chaudhry M, Kumar M, Singhal V, Srinivasan B. Metabolic health tracking using Ultrahuman M1 continuous glucose monitoring platform in non-and pre-diabetic Indians: a multi-armed observational study. Sci Rep. 2024;14(1):6490. PMID:38499685 doi:10.1038/s41598-024-56933-2

36. Caples SM, Rosen CL, Shen WK, et al. The scoring of cardiac events during sleep. J Clin Sleep Med. 2007;3(2):147–154. PMID:17557424

37. Avram R, Tison GH, Aschbacher K, et al. Real-world heart rate norms in the Health eHeart study. NPJ Digit Med. 2019;2:58. PMID:31304404 doi:10.1038/s41746-019-0134-9

38. Huikuri HV, Stein PK. Heart rate variability in risk stratification of cardiac patients. Prog Cardiovasc Dis. 2013;56(2):153–159. PMID:24215747 doi:10.1016/j.pcad.2013.07.003

39. O’Neal WT, Chen LY, Nazarian S, Soliman EZ. Reference ranges for short-term heart rate variability measures in individuals free of cardiovascular disease: the Multi-Ethnic Study of Atherosclerosis (MESA). J Electrocardiol. 2016;49(5):686–690. PMID:27396499 doi:10.1016/j.jelectrocard.2016.06.008

40. Seabold S, Perktold J. Statsmodels: econometric and statistical modeling with Python. In: Proceedings of the 9th Python in Science Conference. 2010:92–96. doi:10.25080/Majora-92bf1922-011

41. Virtanen P, Gommers R, Oliphant TE, et al. SciPy 1.0: fundamental algorithms for scientific computing in Python. Nat Methods. 2020;17(3):261–272. PMID:32015543 doi:10.1038/s41592-019-0686-2

42. Higgins JPT, Thompson SG, Deeks JJ, Altman DG. Measuring inconsistency in meta-analyses. BMJ. 2003;327(7414):557–560. PMID:12958120 doi:10.1136/bmj.327.7414.557

43. World Health Organization. Diabetes, prevalence (%). The Global Health Observatory. https://www.who.int/data/gho/data/indicators/indicator-details/GHO/prevalence-of-diabetes-age-standardized [accessed Oct 13, 2026]

44. NCD Risk Factor Collaboration (NCD-RisC). Worldwide trends in diabetes prevalence and treatment from 1990 to 2022: a pooled analysis of 1108 population-representative studies with 141 million participants. Lancet. 2024;404(10467):2077–2093. PMID:39549716 doi:10.1016/S0140-6736(24)02317-1

45. Summer JV. Normal sleeping heart rate. Sleep Foundation. Updated September 10, 2025. https://www.sleepfoundation.org/physical-health/sleeping-heart-rate [accessed Oct 13, 2026]

46. Cleveland Clinic. Checking your heart rate while sleeping? Here’s what those numbers mean. Cleveland Clinic Health Essentials. January 23, 2024. https://health.clevelandclinic.org/sleeping-heart-rate [accessed Oct 13, 2026]

47. Geovanini GR, Vasques ER, de Oliveira Alvim R, et al. Age and sex differences in heart rate variability and vagal specific patterns -Baependi Heart Study. Glob Heart. 2020;15(1):71. PMID:33150136 doi:10.5334/gh.873

48. Hirshkowitz M, Whiton K, Albert SM, et al. National Sleep Foundation’s updated sleep duration recommendations: final report. Sleep Health. 2015;1(4):233–243. PMID:29073398 doi:10.1016/j.sleh.2015.10.004

49. Shah VN, DuBose SN, Li Z, et al. Continuous glucose monitoring profiles in healthy nondiabetic participants: a multicenter prospective study. J Clin Endocrinol Metab. 2019;104(10):4356–4364. PMID:31127824 doi:10.1210/jc.2018-02763

50. Kianersi S, Wang H, Sofer T, et al. Association between accelerometer-measured irregular sleep duration and type 2 diabetes risk: a prospective cohort study in the UK Biobank. Diabetes Care. 2024;47(9):1647–1655. PMID:39017683 doi:10.2337/dc24-0213

51. Windred DP, Burns AC, Lane JM, et al. Sleep regularity is a stronger predictor of mortality risk than sleep duration: a prospective cohort study. Sleep. 2024;47(1):zsad253. PMID:37738616 doi:10.1093/sleep/zsad253

52. Lloyd-Jones DM, Allen NB, Anderson CAM, et al. Life’s Essential 8: updating and enhancing the American Heart Association’s construct of cardiovascular health: a presidential advisory from the American Heart Association. Circulation. 2022;146(5):e18–e43. PMID:35766027 doi:10.1161/CIR.0000000000001078

53. Dhawale N, Gandhi D, Shanmugam A, et al. Sleep consistency is a low-cost reliable indicator of nocturnal glycemic control: observations from 227,860 nights of real world, free-living smart ring and continuous glucose monitoring data. medRxiv. Preprint posted online March 4, 2026. doi:10.64898/2026.03.04.26347496

54. Anthropic. Claude Opus 5 and Claude Opus 5.5 [large language models]. 2026. https://www.anthropic.com/claude/opus [accessed Oct 13, 2026]

