## Supplemental Information for "Between- and Within-Person Coupling of Nocturnal Heart Rate and Glucose: Retrospective, Real-World Observational Study of 105,196 Nights From 2648 Adults Using Ultrahuman Ring and Continuous Glucose Monitoring"

### Multimedia Appendix 1. Supplementary Information

#### Contents


- Device Validation
  - Comparison With Somfit
  - Comparison With SleepImage
  - Figure S1. Agreement of ring average sleeping heart rate with the Somfit
  - Agreement With the Reference Devices
  - Table S1. Ring continuous overnight heart rate against the SleepImage ring
- Supplementary Figures
  - Figure S2. Region-stratified associations of average sleeping heart rate with the three nocturnal glucose outcomes
  - Figure S3. Association of average sleeping heart rate with each nocturnal glucose outcome, between and within participants
  - Figure S4. Marginal and mutually adjusted associations of average sleeping heart rate with the three nocturnal glucose outcomes
  - Figure S5. Within- and between-participant associations of two secondary autonomic metrics with the three nocturnal glucose outcomes
  - Figure S6. Within- and between-participant associations of nocturnal autonomic metrics with the Night Metabolic Score
  - Figure S7. Correlations among the three sleep-irregularity metrics
  - Figure S8. Marginal and mutually adjusted associations of the three sleep-irregularity metrics with each nocturnal glucose outcome
  - Figure S9. Sleep Score against Night Metabolic Score across participants
- Supplementary Results
  - Robustness of the Heart-Rate Association
  - Regional Differences in the Magnitude of the Coupling
  - Table S2. Cohort composition by geographic region
  - Sensitivity of the Regional Estimates
  - Table S3. Scored sleep duration against the sleep-onset-to-offset interval
  - Table S4. Heart rate and nocturnal mean glucose within body mass index, age, and sex strata
  - Table S5. Sleep-duration irregularity by geographic region
  - Table S6. Measured glycemia against population-representative data, by country

#### Device Validation

The SleepImage comparison [28] and sleep-metric agreement in the Somfit dataset [31] have been reported separately and are summarized here; our own analysis of the Somfit recordings is reported in full. Ethics approval and consent for both comparisons are described in the main text (Ethical Considerations).

##### Comparison With Somfit

Ring sleep metrics and average sleeping heart rate were compared with the Compumedics Somfit, a head-worn device used for medical sleep testing. Five healthy participants (3 male, 2 female; mean age 31 years, standard deviation (SD) 7; self-reported no sleep, metabolic, or cardiovascular conditions) wore the Ultrahuman Ring and the Somfit simultaneously for 32 nights in total, a mean of 6.4 nights each. Volunteers reported no changes in work schedule, diet, medication, or stimulant intake during the study. Only nights where the Somfit rated data quality as good or adequate, with simultaneous ring data logged, were included. Agreement was assessed by comparing medians and coefficients of variation, computing the intraclass correlation coefficient, ICC(3,1), and plotting Bland-Altman differences for sleep duration and average sleeping heart rate. For sleep onset, pairwise between-device differences were tested against zero with a one-sample t test. Outliers, defined as values beyond 1.5 times the interquartile range, were excluded from the comparison statistics. Quality filtering reduced the set to 4 participants (3 male, 1 female) and 23 nights for the sleep-metric comparison; the Bland-Altman comparison of average sleeping heart rate in Figure S1 uses the 21 nights that passed the heart-rate quality filter. The comparison was conducted within a larger study of sleep regularity in which 31 participants wore the ring over 21 days [31]. Sleep-metric agreement from this dataset was subsequently reported in a separate publication by Driller et al [31], who analyzed 20 paired nights. The counts differ because each analysis applies quality criteria to the signals it uses.

##### Comparison With SleepImage

To validate the continuous overnight heart-rate series, ring output was compared with the SleepImage ring, a US Food and Drug Administration (FDA)-cleared finger-worn device used for sleep-apnea testing. Six healthy adults (4 male, 2 female; mean age 31.4 years, SD 6.6; self-reported no sleep, metabolic, or cardiovascular conditions) wore the SleepImage ring and the Ultrahuman Ring simultaneously on the same hand for 6 days in Bangalore, India. Participation was voluntary, and participants were blinded to the analysis. Participants logged perceived sleep disruptions, which were used to screen eligible nights; nights with less than 4 hours of ring-scored sleep or below 70% SleepImage signal quality were excluded. Volunteers reported no changes in work schedule, diet, medication, or stimulant intake. SleepImage data were provided by Empower Sleep with written consent. Agreement was summarized with error metrics and Bland-Altman analysis at the sleep-session level (36 sessions) and at the level of individual heart-rate readings (2349 readings) [28].

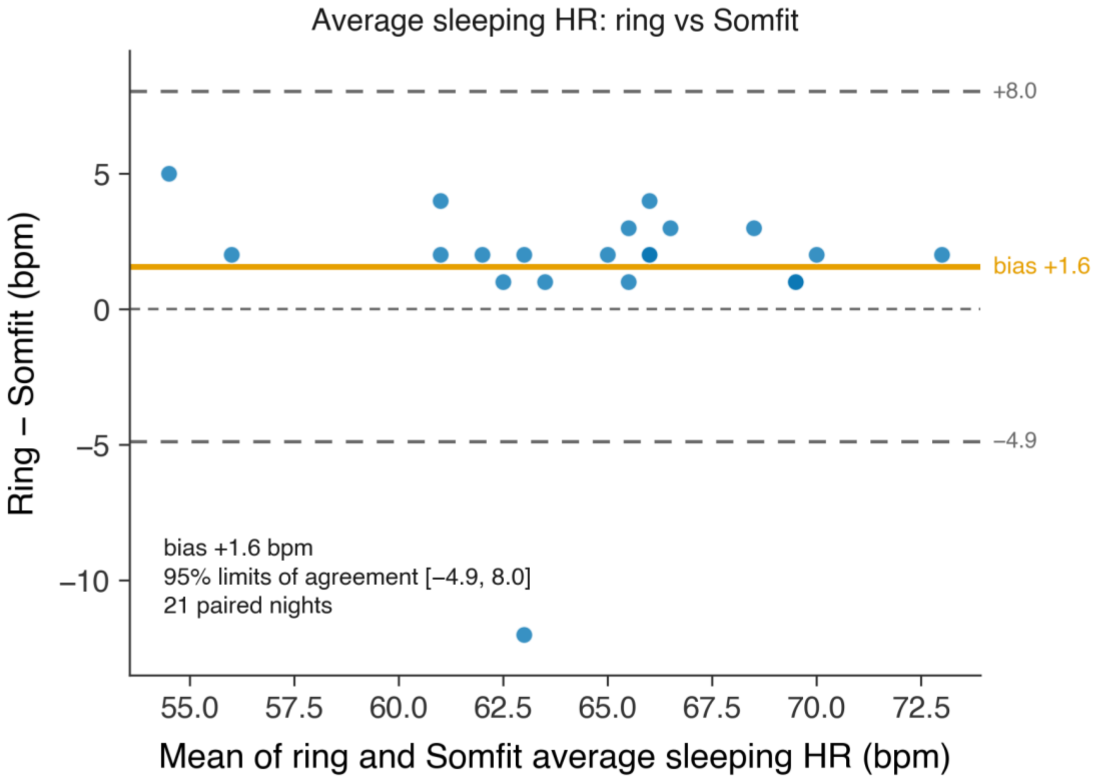


**Figure S1. Agreement of ring average sleeping heart rate with the Somfit.** Bland-Altman plot of the difference between the ring's average sleeping heart rate and the Somfit reference (ring minus Somfit) against the mean of the two, one point per night. The solid orange line is the mean difference, the dashed gray lines the 95% limits of agreement, and the dashed line at zero marks perfect agreement; the bias, limits of agreement, and n are annotated. HR: heart rate.

#### Agreement With the Reference Devices

**Sleep metrics.** In our own analysis of the Somfit dataset, agreement was good for sleep duration (ICC(3,1) 0.87, 95% CI 0.70-0.94) and the between-device difference in sleep onset (3.8 minutes, SD 13.3) did not differ significantly from zero (*P*=.21). Driller et al subsequently analyzed the same dataset and reported concordant values (total sleep time ICC 0.84, r=0.89, bias +1 minute; onset ICC 1.00; offset ICC 0.97) [31].

**Heart rate.** In our own analysis of the Somfit dataset, ring average sleeping heart rate differed from the Somfit reference by a mean of +1.6 bpm, with 95% limits of agreement of -4.9 to +8.0 bpm and an ICC(3,1) of 0.75 (95% CI 0.49-0.89; Figure S1). Continuous overnight heart rate agreed with the FDA-cleared SleepImage ring to a mean absolute error of 2.4 bpm and a root mean square error of 3.9 bpm, with an average difference in readings of -1.2 bpm for the SleepImage-to-Ultrahuman Ring comparison (Table S1) [28].

#### Table S1. Ring continuous overnight heart rate against the SleepImage ring

Agreement of Ultrahuman Ring continuous overnight heart rate with the FDA-cleared SleepImage ring, over 6 participants, 36 sleep sessions, and 2349 paired heart-rate readings [28].

| Reference | Root mean square error (bpm) | Mean absolute error (bpm) | Mean difference (bpm) |
|---|---|---|---|
| SleepImage (FDA-cleared) | 3.9 | 2.4 | -1.2 |

Mean difference is the average difference in readings from the source study's Bland-Altman analysis, for the SleepImage-to-Ultrahuman Ring comparison.

### Supplementary Figures

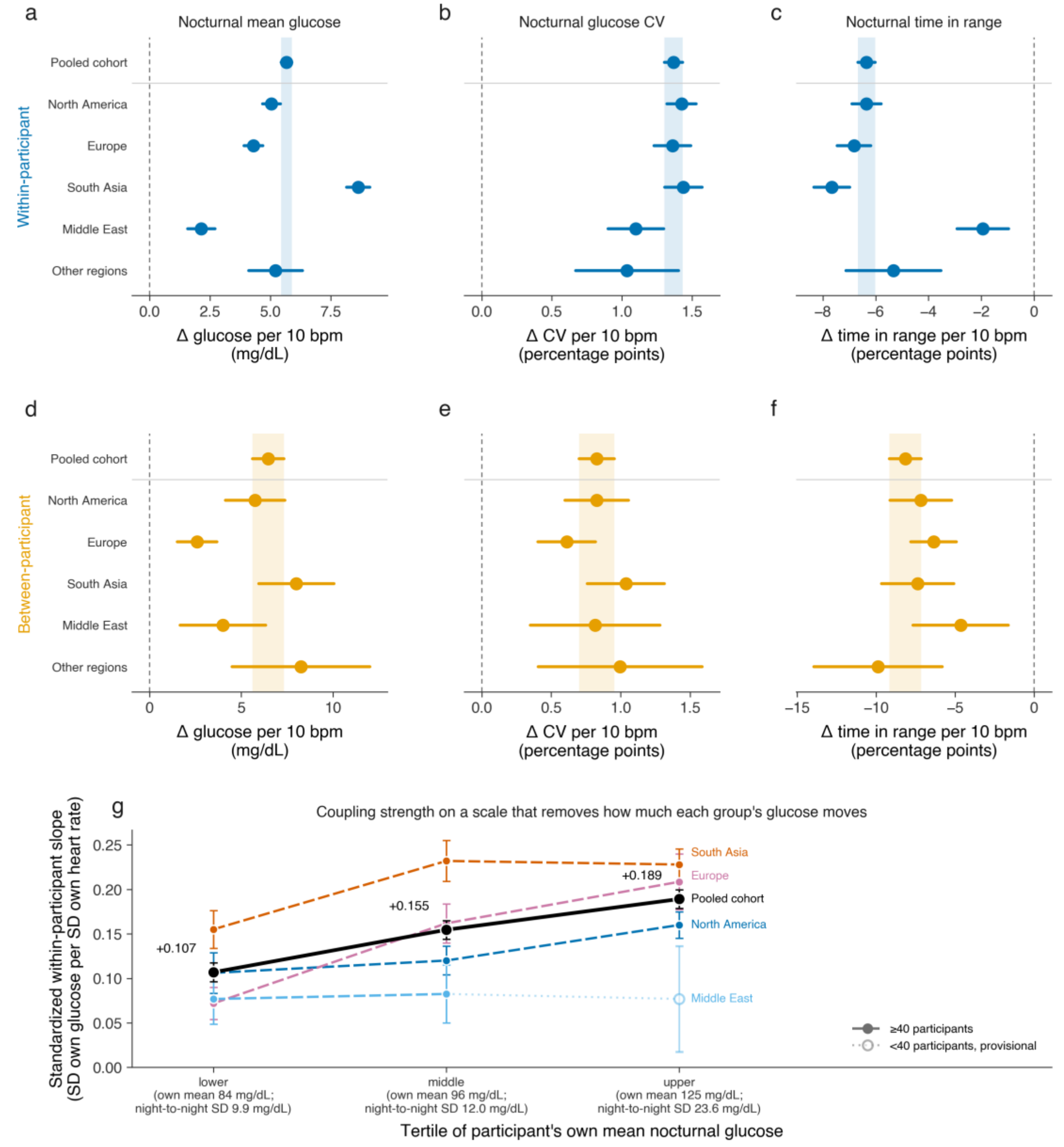


**Figure S2. Region-stratified associations of average sleeping heart rate with the three nocturnal glucose outcomes.** Regions are as defined in Methods (Geographic Region), and the Figure 2 model is refitted within each region. **(a-c)** Within-participant slopes and **(d-f)** between-participant slopes, one column per outcome: mean glucose, glucose coefficient of variation, and time in range, all per 10 bpm. The two levels are shown in separate panels, each against its own pooled interval, because they are on different scales. Points are the adjusted slope, whiskers the 95% confidence interval, the dashed line marks the null, and the shaded vertical band spans the pooled interval for that level only. The pooled cohort is the row above the rule and the region strata are below it; Other regions pools the regions with fewer than 100 participants (n=150 participants). **(g)** The standardized within-participant slope on mean glucose (Methods), refitted inside tertiles of each participant's own mean nocturnal glucose; the night-to-night glucose standard deviation of each tertile is given under its label. Whiskers are 95% confidence intervals, the pooled cohort is the solid black series with its values printed, and each series is directly labeled. The open marker on a dotted line is the provisional Middle East upper-tertile stratum (n=23), excluded from heterogeneity tests (Supplementary Results) (n=2648 participants, 105,196 nights). CV: coefficient of variation; SD: standard deviation.

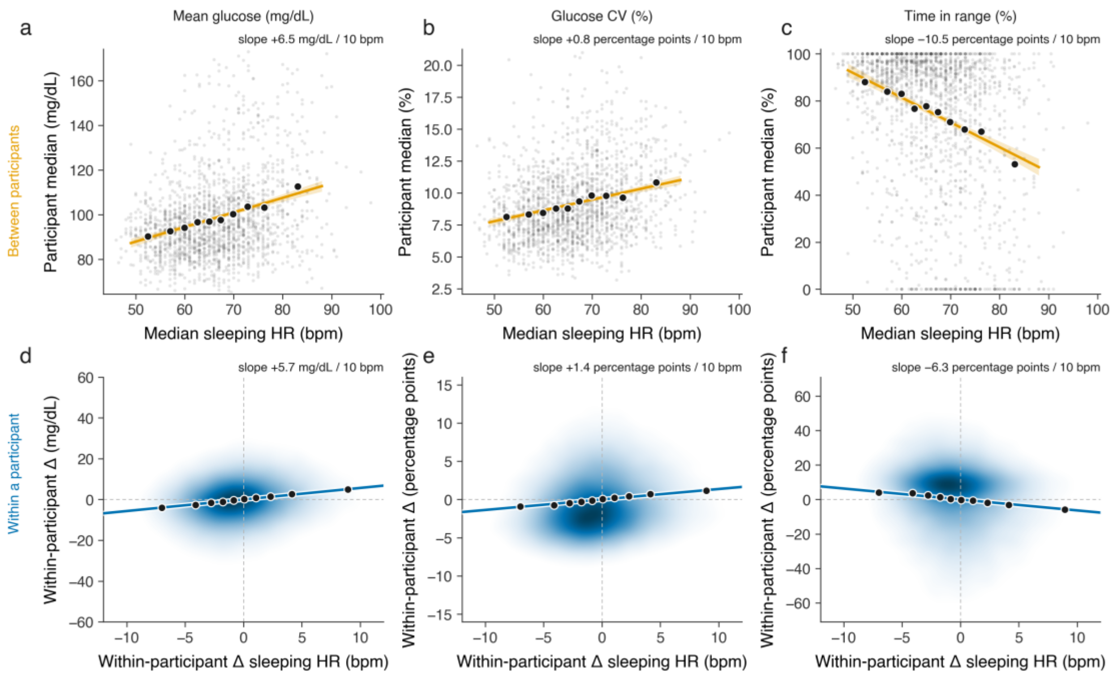


**Figure S3. Association of average sleeping heart rate with each nocturnal glucose outcome, between and within participants.** Each column is one nocturnal outcome (mean glucose, glucose coefficient of variation, time in range). **(a-c)** Between participants: each gray point is one participant's median sleeping heart rate and median outcome over their densest 90-day window containing at least 14 nights; black markers are heart rate decile means and the orange line an ordinary least-squares fit with its 95% confidence band (n=1794 participants). **(d-f)** Within a participant: sleeping heart rate and each outcome are centered on that participant's own mean, shading shows the density of participant-nights, black markers are decile means, and the blue line the pooled within-participant fit (105,196 nights). Both fits are unadjusted least squares, with no age, body mass index, or sex term and no random effect, so their slopes differ from the adjusted mixed-model estimates in Figure 2. The slope per 10 bpm is annotated above each panel. CV: coefficient of variation; HR: heart rate.

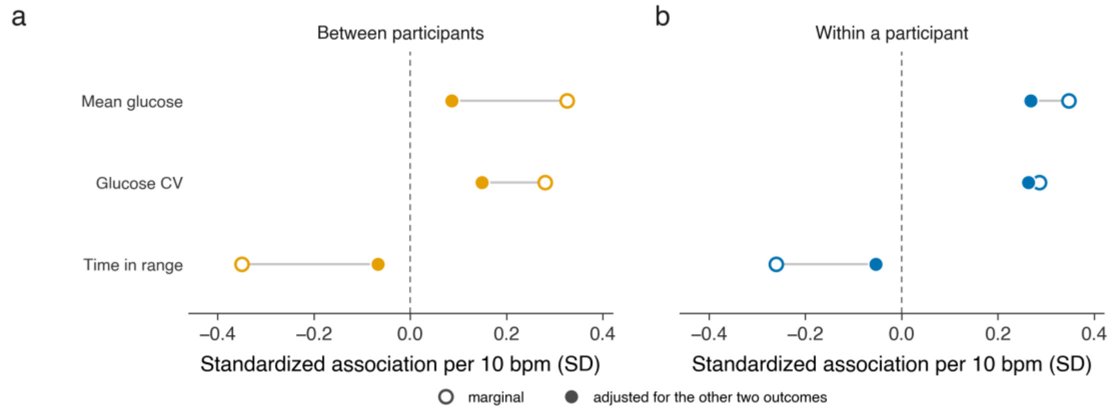


**Figure S4. Marginal and mutually adjusted associations of average sleeping heart rate with the three nocturnal glucose outcomes.** Standardized associations of average sleeping heart rate with mean glucose, glucose coefficient of variation, and time in range, between participants **(a)** and within a participant **(b)**, in outcome standard-deviation units per 10 bpm. Open markers are the marginal association (each outcome regressed on heart rate alone) and filled markers the mutually adjusted association (each outcome additionally adjusted for the other two outcomes), joined by a connecting line; the dashed line marks zero. Estimates are unadjusted least-squares fits: between participants on each participant's median outcomes and median sleeping heart rate, and within participants on nightly deviations from each participant's mean, pooled across nights without a random effect (n=2648 participants, 105,196 nights). CV: coefficient of variation; SD: standard deviation.

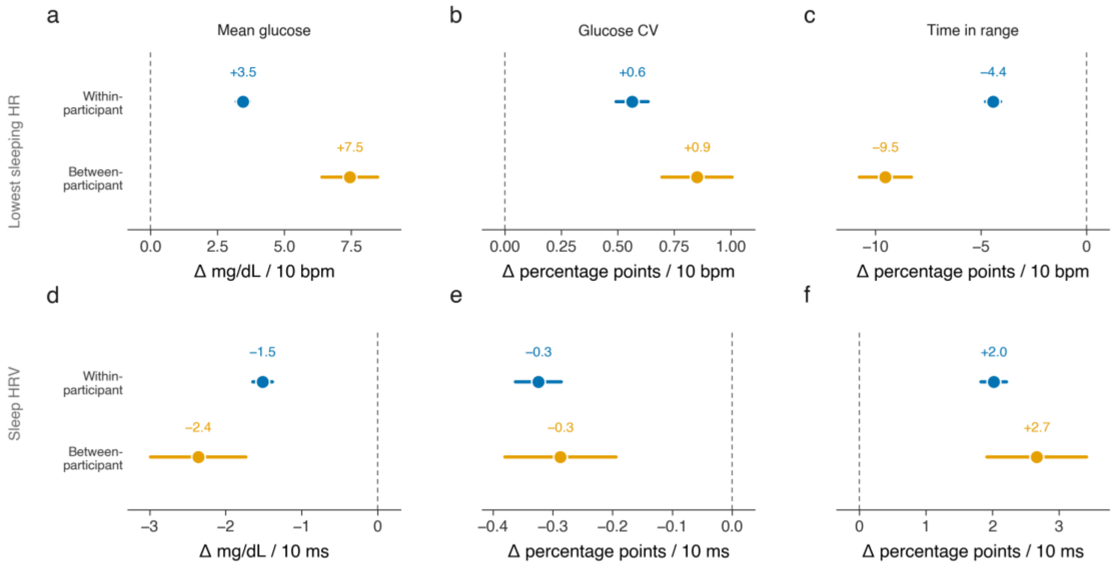


**Figure S5. Within- and between-participant associations of two secondary autonomic metrics with the three nocturnal glucose outcomes.** Decomposition as in Figure 2, for lowest sleeping heart rate (**a-c**, per 10 bpm) and sleep heart rate variability (HRV) measured as the root mean square of successive differences (**d-f**, per 10 ms). Points are adjusted slopes, whiskers are 95% confidence intervals, and the dashed line marks the null (n=2648 participants, 105,196 nights). CV: coefficient of variation; HR: heart rate.

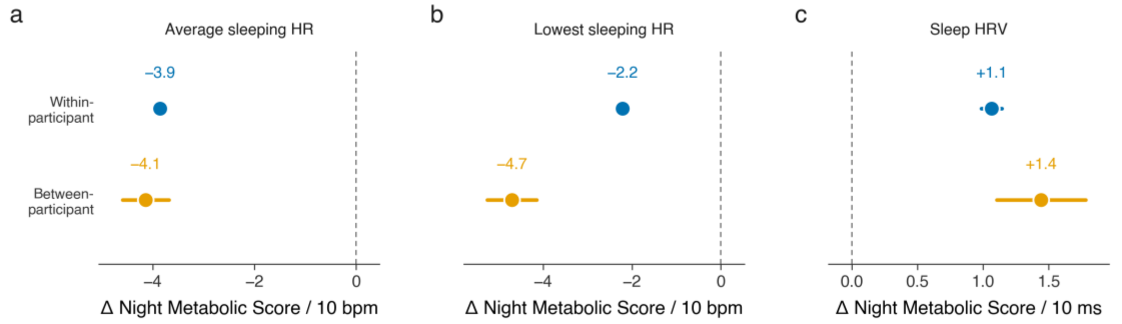


**Figure S6. Within- and between-participant associations of nocturnal autonomic metrics with the Night Metabolic Score.** Decomposition as in Figure 2, with the Night Metabolic Score as the outcome, against average sleeping heart rate **(a)**, lowest sleeping heart rate **(b)**, both per 10 bpm, and sleep heart rate variability (HRV) **(c)**, per 10 ms. Points are adjusted slopes and whiskers 95% confidence intervals. Higher Night Metabolic Score denotes better overnight glucose regulation (n=2648 participants, 105,196 nights). HR: heart rate.

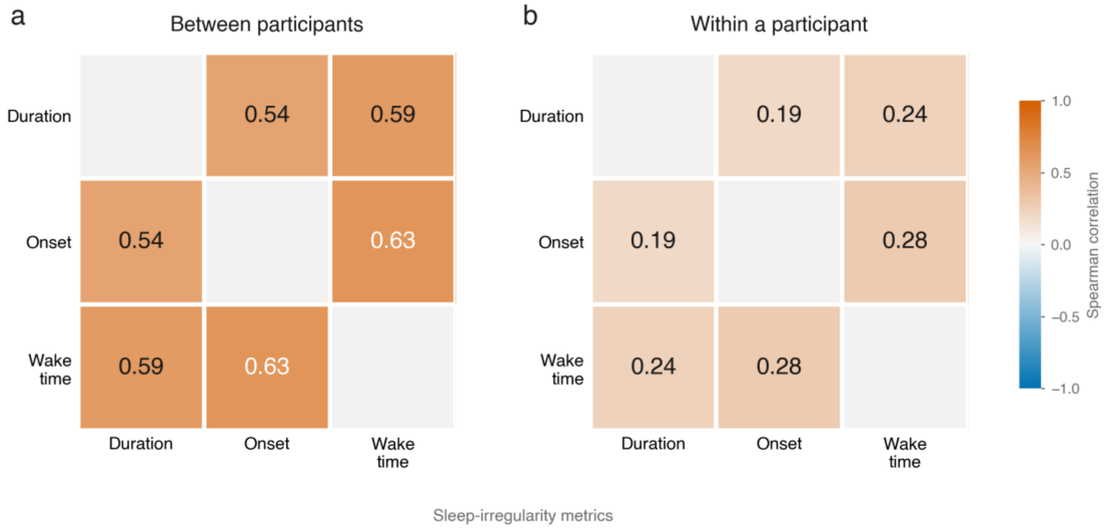


**Figure S7. Correlations among the three sleep-irregularity metrics.** Spearman correlations among night-to-night irregularity in sleep duration, sleep onset, and wake time, computed between participants **(a)**, using each participant's mean irregularity, and within a participant **(b)**, using night-level deviations. Cell color and the printed value give the correlation coefficient (n=2648 participants, 105,196 nights).

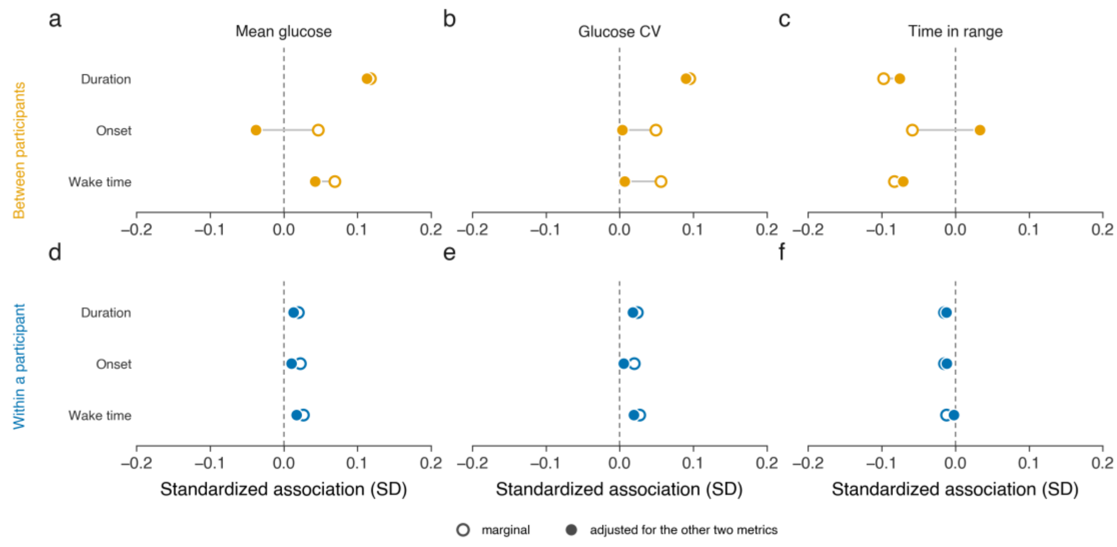


**Figure S8. Marginal and mutually adjusted associations of the three sleep-irregularity metrics with each nocturnal glucose outcome.** Standardized least-squares associations of duration, onset, and wake-time irregularity with each outcome, between participants **(a-c)**, on participant means, and within a participant **(d-f)**, on nightly deviations from the participant's mean, in outcome standard deviations per standard deviation of irregularity; no covariates are included. Open markers are the marginal association and filled markers the association after mutual adjustment for the other two irregularity metrics, joined by a connecting line; the dashed line marks zero. Between participants, under mutual adjustment, duration irregularity remained associated with all three outcomes ($P \leq .001$), wake-time irregularity only with time in range ($P$=.04; $P$=.23 and $P$=.85 otherwise), and onset irregularity with none ($P$=.26, $P$=.91, and $P$=.33). The horizontal scale is common across panels (n=2648 participants, 105,196 nights). CV: coefficient of variation; SD: standard deviation.

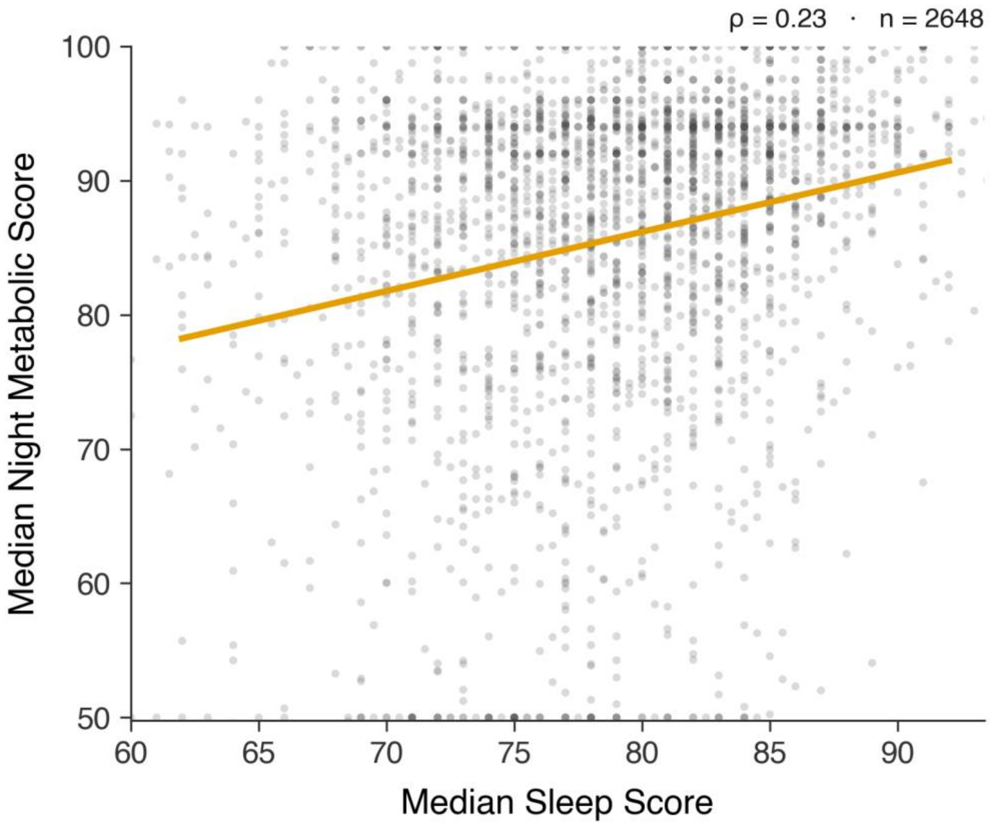


**Figure S9. Sleep Score against Night Metabolic Score across participants.** Each point is one participant's median Sleep Score and median Night Metabolic Score, both platform indices on a 0 to 100 scale where higher denotes better, with an ordinary-least-squares fit (orange). The Spearman correlation (ρ) and n are annotated (n=2648 participants).

#### Supplementary Results

##### Robustness of the Heart-Rate Association

Each robust estimate was compared with a least-squares fit to the same component (Methods). Between participants, those least-squares fits reproduce the mixed-model estimates: +6.43 against +6.47 mg/dL per 10 bpm for mean glucose, +0.81 against +0.83 percentage points for glucose coefficient of variation (CV), and -8.10 against -8.16 percentage points for time in range. Estimators robust to the outcome tail reduced the between-participant slopes by 31% to 38% for mean glucose, 15% to 30% for time in range, and 3% to 9% for glucose CV. Within participants, the time-in-range slope steepened slightly under these estimators, from -6.35 to -6.50 (Huber) and -6.58 (Theil-Sen) percentage points per 10 bpm.

##### Regional Differences in the Magnitude of the Coupling

The association between average sleeping heart rate and nocturnal glucose held in every region, but its magnitude did not (Figure S2). Across the 5 regional strata the within-participant estimates differed for all three outcomes (mean glucose Cochran $Q_4$=329.1, $I^2$=99%; time in range $Q_4$=95.4, $I^2$=96%; glucose CV $Q_4$=12.4, $I^2$=68%), while the between-participant estimates differed only for mean glucose ($Q_4$=29.5, $I^2$=86%, *P*<.001; CV $Q_4$=6.5, $I^2$=39%, *P*=.16; time in range $Q_4$=4.9, $I^2$=18%, *P*=.30). Three factors other than geography contribute to that spread; together they do not account for all of it.

**Scale.** The within-participant standard deviation of nocturnal glucose differed by region: 19.6 mg/dL in South Asia against 16.4 in North America, 13.3 in the Middle East, and 13.1 in Europe. On the standardized scale (Methods), the range of regional slopes narrowed from 4.0-fold to 2.8-fold: +0.076 in the Middle East, +0.134 in North America, +0.140 in Europe, and +0.209 in South Asia, against a pooled +0.155.

**Glycemic case-mix.** The standardized within-participant slope rose 1.8-fold from the lowest to the upper tertile of participants' own mean nocturnal glucose (main text; Figure S2g), so the gradient was not merely a consequence of higher-glucose participants having larger night-to-night swings, although they did: the night-to-night standard deviation of nocturnal glucose was 9.9 mg/dL in the lowest tertile against 23.6 in the upper one. Because the tertiles divide nights rather than participants into thirds, the upper tertile holds 611 participants, and the regions differ substantially in this mix, with 35% of North American and 30% of South Asian participants in the upper tertile against 11% of Middle Eastern and 9% of European ones. Case-mix therefore contributes to the regional spread but does not account for it: differences among the 4 named regions remained significant inside every tertile (lower $Q_3$=37.9, middle $Q_3$=79.9, upper $Q_2$=34.9, all *P*<.001), with South Asia the highest stratum in each.

**Selection.** Selection into the cohort also differs by geography (Table S6).

#### Table S2. Cohort composition by geographic region

Regions are defined in Methods (Geographic Region). Age, body mass index, nocturnal mean glucose, and nights per participant are medians across the participants in each region; the remaining columns are counts of participants, paired nights, and countries, and the percentage male.

| Region | Participants | Nights | Countries | Nights per participant | Age (y) | Body mass index (kg/m$^2$) | Male (%) | Nocturnal mean glucose (mg/dL) |
|---|---|---|---|---|---|---|---|---|
| North America | 796 | 39,478 | 3 | 29 | 44 | 26.5 | 66 | 101 |
| Europe | 783 | 23,983 | 39 | 17 | 42 | 25.9 | 73 | 91 |
| South Asia | 710 | 27,798 | 3 | 22 | 40 | 26.7 | 87 | 98 |

| Region | Participants | Nights | Countries | Nights per participant | Age (y) | Body mass index (kg/m$^2$) | Male (%) | Nocturnal mean glucose (mg/dL) |
|---|---|---|---|---|---|---|---|---|
| Middle East | 209 | 9696 | 9 | 26 | 43 | 26.5 | 73 | 90 |
| Other regions | 150 | 4241 | 33 | 19 | 42 | 25.5 | 75 | 92 |
| **All regions** | **2648** | **105,196** | **87** | **22** | **42** | **26.3** | **75** | **96** |

The Other regions stratum comprises East Asia and Pacific (62 participants), Sub-Saharan Africa (51), Latin America and Caribbean (25), North Africa (9), and Central Asia (3).

**Three of the four primary strata are dominated by a single country** (the United States, India, and the United Arab Emirates; Methods), and their estimates should be read as country rather than regional ones. Europe is the only primary stratum that is geographically diverse, spanning 39 countries, with its largest single country, the United Kingdom, contributing 22% of it.

#### Sensitivity of the Regional Estimates

**Sensitivity to how securely a participant is located.** Restricting the analysis to the 2009 participants who spent at least 80% of their days in their assigned country left four of the five strata within 0.4 mg/dL per 10 bpm of their original within-participant slope on mean glucose (North America +5.1 to +4.7, Europe +4.3 to +3.9, South Asia +8.6 to +8.9, Other regions +5.2 to +5.1) and moved the Middle East from +2.1 to +4.0. That stratum has the lowest location certainty in the cohort, at a median of 77% of days in the assigned country against 100% in North America and South Asia and 93% in Europe, so its distance from the pooled estimate is at least partly a property of the assignment rather than of the participants.

**Ramadan as a potential confounder.** Refitting after removing the 12,408 nights that fell in the two Ramadan periods within the study window left the pooled within-participant slope essentially unchanged, at +5.7 mg/dL per 10 bpm against +5.7, and left every region estimate similarly stable, the largest movement being in the Middle East, from +2.1 to +2.0.

**The provisional Middle East estimate.** Only 23 Middle Eastern participants fall in the upper tertile of the participant's own mean nocturnal glucose, below the 40-participant minimum, because that stratum runs low: its median participant averages 90 mg/dL. The estimate is nevertheless shown in Figure S2g as an open marker so that the series is not silently censored. It is +0.077 (95% CI 0.018-0.136) standard deviations of a participant's own glucose per standard deviation of their own heart rate, close to that region's overall value of +0.076 and below the three regions estimated in the same tertile (+0.160 North

America, +0.209 Europe, +0.228 South Asia). It is excluded from the heterogeneity tests, which use only the strata at or above the minimum.

#### Table S3. Scored sleep duration against the sleep-onset-to-offset interval

Sleep duration in this study is scored sleep, which excludes wake after sleep onset. The alternative measure available from the ring is the interval from sleep onset to sleep offset, the sleep period, which includes wake within the night. The two are not interchangeable.

| Measure | Median per night | Interquartile range |
|---|---|---|
| Scored sleep duration | 7.00 h (420 min) | 367-472 min |
| Sleep onset to offset interval | 7.82 h (469 min) | 414-527 min |
| Difference (interval minus scored sleep) | 40 min | 25-70 min |

The median of the per-night differences is 40 minutes (row above); their mean is 52 minutes, pulled up by a long right tail; and the difference between the two medians, 469 minus 420 minutes, is 49 minutes. The gap corresponds to a median sleep efficiency of 91.3%, and the two measures correlate at r=0.89 across nights. Substituting the interval for scored sleep in the Figure 3 irregularity analysis attenuates the between-participant association of sleep-duration irregularity with nocturnal mean glucose from +4.14 mg/dL per 30 minutes (95% CI 2.63-5.66) to +3.28 (95% CI 2.06-4.50), consistent with the interval carrying night-to-night variation in wake after sleep onset, which scored sleep excludes. Scored sleep is used throughout.

#### Table S4. Heart rate and nocturnal mean glucose within body mass index, age, and sex strata

Slopes of nocturnal mean glucose on average sleeping heart rate, estimated separately inside each stratum with the same person-mean-centered mixed model. Body mass index strata are adjusted for age, body mass index, and sex; age strata are adjusted for body mass index and sex, since age is nearly constant inside each of them; the sex strata are adjusted for age and body mass index, a sex term being unidentifiable inside a single-sex stratum.

| Stratum | Range | Participants | Within-participant slope (mg/dL per 10 bpm) | Between-participant slope |
|---|---|---|---|---|
| Normal weight | 18 to <25 kg/m$^2$ | 987 | +4.8 | +5.0 |
| Overweight | 25 to <30 kg/m$^2$ | 1028 | +5.8 | +7.2 |
| Obese | 30 to 40 kg/m$^2$ | 633 | +6.5 | +7.4 |
| Age 20-29 | 20-29 y | 241 | +5.3 | +3.3 |
| Age 30-39 | 30-39 y | 839 | +5.1 | +4.0 |

| Stratum | Range | Participants | Within-participant slope (mg/dL per 10 bpm) | Between-participant slope |
|---|---|---|---|---|
| Age 40-49 | 40-49 y | 958 | +5.6 | +7.0 |
| Age 50-60 | 50-60 y | 610 | +6.4 | +9.0 |
| Men | | 1975 | +6.2 | +7.6 |
| Women | | 673 | +3.3 | +2.5 |

The within-participant association holds in every stratum of all three variables, so differences in adiposity, age, or sex do not account for it. The between-participant association behaves differently: it is broadly flat across body mass index but rises steeply with age, from +3.3 mg/dL per 10 bpm in the youngest decade to +9.0 in the oldest. A between-participant estimate compares different people and can accumulate anything that tracks age, whereas the within-participant estimate compares a participant with themselves and cannot.

**Age is not a confounder of the within-participant estimate, by construction.** Person-mean centering removes every characteristic that is constant within a participant, and age is effectively constant over a recording window whose median length is 66 days. Refitting the primary model with no age term, with linear age, with quadratic and cubic age, and with age as a four-level factor returned a within-participant slope of +5.661 mg/dL per 10 bpm in every case, identical to three decimal places. The between-participant slope did move, from +6.72 with no age term to +6.47 with linear age, and allowing curvature changed it no further: the quadratic, cubic, and factor forms gave +6.47, +6.45, and +6.47, so a linear age term is sufficient for these estimates. Age itself was associated with +3.0 mg/dL of nocturnal glucose per decade.

**Sex is a negative confounder, and also modifies the association.** Women in this cohort had higher average sleeping heart rate than men, 68.1 against 65.5 bpm, but lower nocturnal glucose, 91.4 against 98.9 mg/dL, so sex pushes the two variables in opposite directions and omitting it biases the between-participant estimate downward (main text). The within-participant estimate was unmoved, at +5.661 with and without the term.

Scale does not fully explain the sex difference in the within-participant slope (+6.2 in men against +3.3 mg/dL per 10 bpm in women; main text). Glucose moves less from night to night in women, with a within-participant standard deviation of 13.1 mg/dL against 17.1 in men, and so does heart rate, at 4.1 against 4.6 bpm; rescaling each sex's slope by both standard deviations leaves a 1.6-fold difference in place of the raw 1.9-fold. Adiposity does not explain it either, since restricting both sexes to the normal-weight stratum widens rather than narrows the gap, to +5.7 against +2.4. The mechanism cannot be identified from these data (Discussion).

**Drift over a participant's own recording window.** Nocturnal glucose declined slightly within participants across their wear period, by 0.36 mg/dL per month. Adding a within-participant time term to the primary model moved the within-participant slope from +5.66 to +5.87 and the between-participant slope from +6.47 to +6.06. Neither changes any conclusion, and the within-participant association is slightly stronger once this drift is removed.

#### Table S5. Sleep-duration irregularity by geographic region

Between-participant slope of each nocturnal glucose outcome on night-to-night sleep-duration irregularity, per 30 minutes, from the same person-mean-centered mixed model fitted separately inside each region. Values are the slope with its 95% confidence interval. This is the secondary finding of the main text, and it is a between-participant association, so each region estimate rests on the number of participants in that region rather than on the number of nights.

| Region | Mean glucose (mg/dL) | Glucose CV (percentage points) | Time in range (percentage points) |
|---|---|---|---|
| Pooled cohort | +4.14 (2.63-5.66) | +0.52 (0.30-0.75) | -3.99 (-5.80 to -2.18) |
| North America | +3.53 (0.62-6.44) | +0.94 (0.53-1.35) | -4.08 (-7.60 to -0.56) |
| Europe | +1.68 (-0.10 to 3.46) | +0.35 (0.002-0.69) | -2.95 (-5.41 to -0.50) |
| South Asia | +6.05 (2.39-9.71) | +1.09 (0.59-1.58) | -6.32 (-10.35 to -2.29) |
| Middle East | +1.75 (-2.06 to 5.56) | +0.08 (-0.69 to 0.85) | -0.35 (-5.27 to 4.57) |
| Other regions | -0.92 (-6.98 to 5.15) | -0.65 (-1.59 to 0.29) | +2.44 (-4.22 to 9.10) |

The direction of the sleep-duration association is preserved in all four primary regions for all three outcomes, and the interval excludes the null in North America and South Asia for all three and in Europe for two. All three Middle East intervals include the null, and the estimates for Other regions (150 participants) reverse direction on all three outcomes with intervals that also include the null. The European estimate for glucose CV only just excludes the null ($P$=.049). Across the five strata, heterogeneity is detected only for glucose CV (Cochran $Q_4$=17.1, $P$=.002; mean glucose $Q_4$=6.3, $P$=.18; time in range $Q_4$=6.6, $P$=.16). The sleep-duration finding is therefore consistent in direction across regions but is not independently established within each one, and the region-level estimates should not be read as separate results.

#### Table S6. Measured glycemia against population-representative data, by country

We are not aware of a population-representative continuous-monitoring dataset for any of these countries, so the reference is age-standardized diabetes prevalence from population-representative surveys [43,44], compared on rank rather than level because the two are different quantities. National rates are weighted by the sex mix of our own participants in that country. Countries contributing fewer than 40 participants are not compared; the eight below hold 80% of the cohort. Glucose values are means of participant means.

| Country | Our participants | Our 24-h mean glucose (mg/dL) | Our nocturnal mean glucose (mg/dL) | National diabetes prevalence (%) |
|---|---|---|---|---|
| United Arab Emirates | 171 | 96.9 | 90.9 | 26.0 |
| India | 707 | 106.8 | 98.7 | 21.7 |
| United States | 781 | 109.9 | 104.3 | 14.0 |
| Czechia | 122 | 98.8 | 90.7 | 11.2 |
| United Kingdom | 176 | 97.5 | 91.5 | 9.8 |
| Germany | 67 | 95.9 | 89.8 | 7.3 |
| Switzerland | 45 | 96.0 | 88.1 | 4.7 |
| France | 44 | 98.3 | 91.6 | 4.3 |

The ordering diverges most for the United Arab Emirates (main text). In a leave-one-country-out check, removing the United Arab Emirates moved the rank correlation from 0.40 to 0.71, a larger shift than removing any other country (next largest, France, 0.57; the remaining six moved it to between 0.29 and 0.39).

**Absolute plausibility.** Rank aside, the measured levels can be checked for plausibility against published continuous-monitoring reference cohorts.

| Cohort | 24-hour mean glucose (mg/dL) |
|---|---|
| Screened healthy individuals, United States (n=153, ages 7 to 80) [49] | 98.5 |
| Screened healthy adults, India (n=53, ages 25 to 50) [35] | 102.4 |
| Adults with prediabetes, India (n=52, ages 25 to 50) [35] | 112.2 |
| This cohort, all participants | 103.8 |
| This cohort, North America | 109.8 |
| This cohort, South Asia | 106.8 |
| This cohort, Europe | 97.3 |
| This cohort, Middle East | 97.1 |

The North American and South Asian strata sit above both screened healthy values and below the prediabetes value, while the European and Middle Eastern strata sit below the screened healthy values. The Middle Eastern stratum contrasts most sharply with the population data: its measured mean is the lowest of the four strata, while the United Arab Emirates, the only one of the eight countries that falls in that region, has the highest national diabetes prevalence. Taken with the rank comparison, this is consistent with selection into the cohort differing by geography.

#### References


28. Krishnan N, et al. Sleep heart rate sensing by Ultrahuman Ring AIR. Ultrahuman Science Portal. 2026. https://science.ultrahuman.com/studies/sleep-heart-rate-sensing
29. Driller M, Bodner ME, Fenuta A, Stevenson S, Suppiah H. How does the Sleep Regularity Questionnaire relate to wearable- and diary-derived sleep regularity? J Circadian Rhythms. 2026;24:2. PMID:42145918 doi:10.5334/jcr.265
30. Chaudhry M, Kumar M, Singhal V, Srinivasan B. Metabolic health tracking using Ultrahuman M1 continuous glucose monitoring platform in non- and pre-diabetic Indians: a multi-armed observational study. Sci Rep. 2024;14(1):6490. PMID:38499685 doi:10.1038/s41598-024-56933-2
31. World Health Organization. Diabetes, prevalence (%). The Global Health Observatory. https://www.who.int/data/gho/data/indicators/indicator-details/GHO/prevalence-of-diabetes-age-standardized [accessed Oct 13, 2026]
32. NCD Risk Factor Collaboration (NCD-RisC). Worldwide trends in diabetes prevalence and treatment from 1990 to 2022: a pooled analysis of 1108 population-representative studies with 141 million participants. Lancet. 2024;404(10467):2077-2093. PMID:39549716 doi:10.1016/S0140-6736(24)02317-1
33. Shah VN, DuBose SN, Li Z, et al. Continuous glucose monitoring profiles in healthy nondiabetic participants: a multicenter prospective study. J Clin Endocrinol Metab. 2019;104(10):4356-4364. PMID:31127824 doi:10.1210/jc.2018-02763
