## Supplemental Information STROBE Checklist for "Between- and Within-Person Coupling of Nocturnal Heart Rate and Glucose: Retrospective, Real-World Observational Study of 105,196 Nights From 2648 Adults Using Ultrahuman Ring and Continuous Glucose Monitoring"

### Multimedia Appendix 2. STROBE Statement: Checklist of Items for Cohort Studies

Page and line numbers refer to the line-numbered main text. Locations also name the section heading in the main text (Methods, Results, Discussion) or Multimedia Appendix 1.

| Item | No. | Recommendation | Page, lines | Where and how reported |
|---|---|---|---|---|
| **Title and abstract** | 1 | (a) Indicate the study's design with a commonly used term in the title or the abstract | p 1, lines 1-5; p 1, line 27 | Title ("Retrospective, Real-World Observational Study"); Abstract, Methods ("retrospective observational study") |
| | | (b) Provide in the abstract an informative and balanced summary of what was done and what was found | pp 1-2, lines 18-56 | Abstract |
| **Introduction** | | | | |
| Background/ rationale | 2 | Explain the scientific background and rationale for the investigation being reported | pp 2-3, lines 57-105 | Introduction |
| Objectives | 3 | State specific objectives, including any prespecified hypotheses | p 3, lines 97-105 | Introduction, final paragraph (the objectives are stated as two questions, each with the expected direction of the association); Abstract, Objective |
| **Methods** | | | | |
| Study design | 4 | Present key elements of study design early in the paper | p 3, line 108 | Methods: Study Design and Participants (retrospective observational study of concurrent |

| Item | No. | Recommendation | Page, lines | Where and how reported |
|---|---|---|---|---|
| | | | | ring and continuous glucose monitor users) |
| Setting | 5 | Describe the setting, locations, and relevant dates, including periods of recruitment, exposure, follow-up, and data collection | p 3, lines 109-111; pp 9-10, lines 293-341 | Methods: Study Design and Participants (January 2024 to June 2025, free-living data from the Ultrahuman platform); Methods: Geographic Region (country assignment and World Bank regions); Results: Participant Characteristics (87 countries) |
| Participants | 6 | (a) Give the eligibility criteria, and the sources and methods of selection of participants. Describe methods of follow-up | pp 3-4, lines 107-114; pp 6-8, lines 205-240 | Methods: Study Design and Participants; Methods: Analysis Dataset and Inclusion Criteria; Figure 1. Participants were all concurrent users of both devices in the study window who met the inclusion criteria; follow-up was the set of nights on which both devices were worn |
| | | (b) For matched studies, give matching criteria and number of exposed and unexposed | Not applicable | Not applicable: no matching |
| Variables | 7 | Clearly define all outcomes, exposures, predictors, potential confounders, and effect modifiers. Give diagnostic criteria, if applicable | pp 4-6, lines 134-204; pp 8-11, lines 241-380 | Methods: Devices and Measurements (exposure: average sleeping heart rate; outcomes: nocturnal mean glucose, glucose coefficient of variation, and time in range); Methods: Statistical Analysis (sleep irregularity predictors; |

| Item | No. | Recommendation | Page, lines | Where and how reported |
| --- | --- | --- | --- | --- |
| | | | | confounders age, body mass index, and sex; effect modification by region and glucose tertile); Table S4 in Multimedia Appendix 1 (body mass index, age, and sex strata) |
| Data sources/ measurement | 8 | For each variable of interest, give sources of data and details of methods of assessment (measurement). Describe comparability of assessment methods if there is more than one group | pp 4-6, lines 134-204 | Methods: Devices and Measurements (Ultrahuman Ring; Validation of Ring Sleep and Heart-Rate Measures; Continuous Glucose Monitoring and Nocturnal Glucose Metrics; Platform Composite Scores); Multimedia Appendix 1: Device Validation. The same ring and glucose-metric processing were used for every participant; CGM data from 3 FreeStyle Libre models were resampled to a common 15-minute interval |
| Bias | 9 | Describe any efforts to address potential sources of bias | pp 6-8, lines 205-240; p 8, lines 250-275; pp 9-10, lines 293-341; p 11, lines 355-372; pp 19-20, lines 595-605 | Methods: Analysis Dataset and Inclusion Criteria (filters for measurement artifact); Methods: Within-Participant and Between-Participant Decomposition (person-mean centering removes time-invariant confounding); Methods: Robustness; Methods: Geographic Region (location and Ramadan sensitivity analyses); Discussion (confounding, selection, and measurement) |

| Item | No. | Recommendation | Page, lines | Where and how reported |
|---|---|---|---|---|
| Study size | 10 | Explain how the study size was arrived at | p 3, lines 109-111 | Methods: Study Design and Participants; Figure 1. All concurrent users in the study window were considered (5859 participants, 227,860 nights), and the analysis cohort is those meeting the inclusion criteria. No a priori sample size calculation was made for this retrospective analysis |
| Quantitative variables | 11 | Explain how quantitative variables were handled in the analyses. If applicable, describe which groupings were chosen and why | p 7, line 228; p 8, lines 261-262; p 8, lines 262-265; p 10, lines 324-325 | Methods: Statistical Analysis (heart rate per 10 bpm, sleep irregularity per 30 minutes, heart rate variability per 10 ms; within- and between-participant components; body mass index, age, and sex strata); Methods: Geographic Region (tertiles of participant mean nocturnal glucose, with cut-points); Methods: Analysis Dataset and Inclusion Criteria (quartiles of Night Metabolic Score, with cut-points); Table S4 in Multimedia Appendix 1 |
| Statistical methods | 12 | (a) Describe all statistical methods, including those used to control for confounding | pp 8-11, lines 241-380 | Methods: Statistical Analysis (person-mean-centered linear mixed models with a random intercept for participant, adjusted for age, body mass index, and sex) |
| | | (b) Describe any methods used to examine subgroups and | p 8, lines 262-265; pp 9-10, lines 293- | Methods: Statistical Analysis (body mass index, age, and sex strata; sex |

| Item | No. | Recommendation | Page, lines | Where and how reported |
|---|---|---|---|---|
| | | interactions | 341; p 10, lines 342-354 | interaction terms); Methods: Geographic Region; Methods: Metabolic Extreme Groups and Per-Participant Coupling; Table S4 in Multimedia Appendix 1; Multimedia Appendix 1, Supplementary Results |
| | | (c) Explain how missing data were addressed | p 5, lines 164-167; p 7, lines 220-222 | Methods: Continuous Glucose Monitoring and Nocturnal Glucose Metrics (glucose gaps longer than 32 minutes treated as missing; sessions below 80% data density excluded); Methods: Analysis Dataset and Inclusion Criteria (minimum of 10 valid paired nights per participant to allow for gaps in wear). Glucose gaps of up to 32 minutes were bridged by linear interpolation during resampling; otherwise, nights and sessions with missing data were excluded, and no analysis variable was imputed |
| | | (d) If applicable, explain how loss to follow-up was addressed | Not applicable | Not applicable: retrospective analysis of recorded nights, with no scheduled follow-up |
| | | (e) Describe any sensitivity analyses | p 10, lines 327-332; p 11, lines 355-372 | Methods: Robustness; Methods: Geographic Region (location-certainty and Ramadan sensitivity analyses); Multimedia Appendix 1, Supplementary Results |

| Item | No. | Recommendation | Page, lines | Where and how reported |
|---|---|---|---|---|
| **Results** | | | | |
| Participants | 13 | (a) Report numbers of individuals at each stage of study—eg numbers potentially eligible, examined for eligibility, confirmed eligible, included in the study, completing follow-up, and analysed | p 7, line 223; p 7, line 225; p 7, lines 234-238 | Figure 1 (source records, 5859 participants and 227,860 nights; analysis cohort, 2648 participants and 105,196 nights; subcohorts); Methods: Analysis Dataset and Inclusion Criteria (the glucose bound and re-evaluated night minimum removed 419 nights and 3 participants) |
| | | (b) Give reasons for non-participation at each stage | pp 6-8, lines 205-240 | Methods: Analysis Dataset and Inclusion Criteria; Figure 1 (inclusion criteria listed in the order applied) |
| | | (c) Consider use of a flow diagram | p 7, lines 234-238 | Figure 1 |
| Descriptive data | 14 | (a) Give characteristics of study participants (eg demographic, clinical, social) and information on exposures and potential confounders | pp 11-14, lines 383-401 | Results: Participant Characteristics; Table 1; Table S2 in Multimedia Appendix 1 (by region) |
| | | (b) Indicate number of participants with missing data for each variable of interest | Not applicable | Not applicable: nights with incomplete data were excluded before analysis (item 12c) |
| | | (c) Summarise follow-up time (eg, average and total amount) | p 11, line 385 | Results: Participant Characteristics (median 22 nights per participant, IQR 13-44; 105,196 nights in total); Multimedia Appendix 1, Supplementary Results (median |

| Item | No. | Recommendation | Page, lines | Where and how reported |
| --- | --- | --- | --- | --- |
| | | | | recording window 66 days) |
| Outcome data | 15 | Report numbers of outcome events or summary measures over time | pp 11-14, lines 383-401 | Results: Participant Characteristics; Table 1 (the outcomes are continuous nightly glucose measures, reported as medians and interquartile ranges) |
| Main results | 16 | (a) Give unadjusted estimates and, if applicable, confounder-adjusted estimates and their precision (eg, 95% confidence interval). Make clear which confounders were adjusted for and why they were included | pp 14-15, lines 402-448 | Results: Sleeping Heart Rate and Nocturnal Glucose; Figure 2 (adjusted for age, body mass index, and sex, with 95% CIs); Figure S3 in Multimedia Appendix 1 (unadjusted estimates); Multimedia Appendix 1, Supplementary Results (effect of the age and sex terms) |
| | | (b) Report category boundaries when continuous variables were categorized | p 7, line 228; p 8, lines 262-265; p 10, lines 324-325 | Methods: Statistical Analysis (body mass index 18 to <25, 25 to <30, and 30 to 40 kg/m$^2$; age by decade); Methods: Geographic Region (glucose tertile cut-points); Methods: Analysis Dataset and Inclusion Criteria (Night Metabolic Score quartile cut-points); Table S4 in Multimedia Appendix 1 |
| | | (c) If relevant, consider translating estimates of relative risk into absolute risk for a meaningful time period | Not applicable | Not applicable: continuous outcomes, no relative risks |
| Other analyses | 17 | Report other analyses done—eg analyses of subgroups and | p 15, lines 449-461; pp 15-16, lines 462- | Results: Redundancy Among the Glucose Outcomes; Alternative |

| Item | No. | Recommendation | Page, lines | Where and how reported |
|---|---|---|---|---|
| | | interactions, and sensitivity analyses | 469; pp 16-17, lines 470-503; pp 17-18, lines 504-534; pp 18-19, lines 535-561 | Autonomic Metrics; Night-to-Night Sleep Irregularity; Consistency Across Geographic Regions; Metabolic Extreme Groups and Per-Participant Coupling; Multimedia Appendix 1, Supplementary Results |
| **Discussion** | | | | |
| Key results | 18 | Summarise key results with reference to study objectives | p 19, lines 563-574 | Discussion, first paragraph |
| Limitations | 19 | Discuss limitations of the study, taking into account sources of potential bias or imprecision. Discuss both direction and magnitude of any potential bias | pp 19-20, lines 595-605; p 21, lines 647-664 | Discussion (time-varying confounding, selection of device users, sex differences, measurement considerations, and outlier fences on a glucose-derived score) |
| Interpretation | 20 | Give a cautious overall interpretation of results considering objectives, limitations, multiplicity of analyses, results from similar studies, and other relevant evidence | p 11, lines 373-380; pp 19-21, lines 562-676 | Discussion; Conclusions; Methods: Multiplicity and Joint Testing |
| Generalisability | 21 | Discuss the generalisability (external validity) of the study results | p 20, lines 606-613 | Discussion (regional strata describe device users rather than national populations; self-selected consumers; cohort glucose relative to published reference cohorts); Table S6 in Multimedia Appendix 1 |
| **Other information** | | | | |

| Item | No. | Recommendation | Page, lines | Where and how reported |
| --- | --- | --- | --- | --- |
| Funding | 22 | Give the source of funding and the role of the funders for the present study and, if applicable, for the original study on which the present article is based | p 22, lines 688-691 | Funding (Ultrahuman internal research funds; apart from the work of the authors it employed, the funder had no role in the study design, analysis, interpretation, or decision to publish); Conflicts of Interest |

Checklist items are from the STROBE Statement for cohort studies (von Elm E, Altman DG, Egger M, et al. The Strengthening the Reporting of Observational Studies in Epidemiology (STROBE) statement: guidelines for reporting observational studies. PLoS Med. 2007;4(10):e296), which is distributed under a Creative Commons Attribution license.
